# Data-driven analysis shows robust links between fatigue and depression in early multiple sclerosis

**DOI:** 10.1101/2022.01.13.22269128

**Authors:** Yuan-Ting Chang, Patrick K.A. Kearns, Alan Carson, David C. Gillespie, Rozanna Meijboom, Agniete Kampaite, Maria del C. Valdés Hernández, Christine Weaver, Amy Stenson, Niall MacDougall, Jonathan O’Riordan, Margaret Ann Macleod, Francisco Javier Carod-Artal, Peter Connick, Adam D. Waldman, Siddharthan Chandran, Peter Foley

## Abstract

Fatigue is common and disabling in multiple sclerosis, yet its mechanisms are poorly understood. In particular, overlap in measures of fatigue and depression complicates interpretation. A clearer understanding of relationships between fatigue and key clinical, neuropsychiatric and imaging variables including depression could yield clinically relevant mechanistic insight. We applied a data-driven multivariate network approach to quantify relationships between fatigue and other variables in early multiple sclerosis.

Data were collected from Scottish patients with newly diagnosed, immunotherapy-naïve, relapsing-remitting multiple sclerosis at baseline and month 12 follow-up in FutureMS, a nationally representative multicentre cohort. Subjective fatigue was assessed using the validated Fatigue Severity Scale. Detailed phenotyping included measures assessing physical disability, affective disorders, objective cognitive performance, subjective sleep quality, and structural brain imaging. Bivariate correlations between fatigue and other variables were calculated. Network analysis was then conducted to estimate partial correlations between variables, after accounting for all other included variables. Secondary networks included individual depressive symptoms, to control for overlapping symptom items in measures of fatigue and depression.

Data from 322 participants at baseline, and 323 at month 12, were included. At baseline, 49.5% of the cohort reported clinically significant fatigue. Bivariate correlations confirmed that fatigue severity was significantly correlated with all included measures of physical disability, affective disturbance (anxiety and depression), cognitive performance (processing speed and memory/attention), and sleep quality, but not with structural brain imaging variables including normalized lesion and grey matter volumes. In the network analysis, fatigue showed strong correlations with depression, followed by Expanded Disability Status Scale. Weak connections with walking speed, subjective sleep quality and anxiety were identified. After separately controlling for measurement of “tiredness” in our measure of depression, some key depressive symptoms (anhedonia, subjective concentration deficits, subjectively altered speed of movement, and appetite) remained linked to fatigue. Conversely, fatigue was not linked to objective cognitive performance, white matter lesion volume, or grey matter volumes (cortical, subcortical or thalamic). Results were consistent at baseline and month 12. Depression was identified as the most central variable in the networks. Correlation stability coefficients and bootstrapped confidence intervals of the edge weights supported stability of the estimated networks.

Our findings support robust links between subjective fatigue and depression in early relapsing-remitting multiple sclerosis, despite absence of links between fatigue and either objective cognitive performance, or structural brain imaging variables. Depression, including specific depressive symptoms, could be a key target of treatment and research in multiple sclerosis-related fatigue.

## Introduction

Multiple sclerosis is an autoimmune-mediated neuroinflammatory and neurodegenerative condition which is a major cause of morbidity in young adults and affects over two million people worldwide.^1^ Fatigue is frequently described by pwMS as one of their most disabling symptoms,^2,3^ with the estimated prevalence ranging from 38% to 83%.^2^ Fatigue can be defined as “a subjective lack of physical and/or mental energy that is perceived by the individual or the care-giver to interfere with usual or desired activity”.^4^ Although fatigue is distinct from physical fatigability, depression, tiredness, and other phenomena which are frequently present in people with multiple sclerosis (pwMS),^2,5^ clearly differentiating subjective fatigue from these phenomena can be challenging.

Despite the high prevalence, treatment options for fatigue in pwMS are of limited effectiveness.^6,7^ This in part reflects limited mechanistic understanding and an absence of clear biomarkers.^8–11^ A fundamental challenge to the development of such mechanistic understanding is the overlap and interrelatedness with other common symptoms. Multiple factors have been linked to fatigue in pwMS, including physical disability,^11^ obesity,^12^ depression,^11^ anxiety,^10^ sleep disturbance,^13^ and subjective cognitive difficulty.^14^ However, studies to date have highlighted inconsistency in many of these associations. For example, reported links between subjective fatigue and physical disability have been diminished or removed in some studies by correction for depression as a confounding variable.^15^ Similarly, subjective cognitive difficulty in pwMS experiencing fatigue has been reported in the absence of detectable cognitive impairment on neuropsychological testing.^14^ Reported imaging correlates of fatigue are also inconsistent, with some studies reporting no significant link between fatigue and imaging outcomes (including T2 lesion volume and whole brain volume)^10^ but others reporting focal atrophy and altered functional connectivity.^16^ Given the clinical complexity, multivariate analysis that simultaneously evaluates physical, cognitive, affective, sleep, and imaging variables, is likely necessary to an improved understanding of the complex inter-relationships between these variables, and ultimately to advance understanding of pathogenic mechanisms.

We therefore aimed to examine the complex relationships between fatigue, physical, affective, and cognitive functions in pwMS using a network analysis approach. Network analysis is based on the conceptualisation that symptoms or variables in the network can act as active agents to mutually reinforce each other rather than correlate only through a common latent cause.^17,18^ We hypothesised that fatigue in pwMS together with other physical, affective, sleep and cognitive symptoms play active roles to influence each other in a network system. Imaging variables representing the objective changes in brain structures might also provide insight with respect to the pathophysiology of fatigue in such a network. The use of network analysis (including a range of clinical, imaging and other variables) allows for both the quantification and visualisation of relationships between fatigue and other network nodes, and between other nodes apart from fatigue. In addition, network centrality indices may be used to infer the relative influence of the included nodes in the estimated networks.

We quantified the relationships between subjective fatigue and variables reflecting aspects of physical disability, cognitive performance, depression, anxiety, sleep quality, and MRI brain imaging metrics, deploying mixed graphical model networks at study baseline and one-year follow up to examine and visualise the correlates of fatigue in pwMS.

## Materials and Methods

### Participants

The study participants were drawn from FutureMS,^19^ a nationally representative cohort of patients with newly diagnosed relapsing remitting MS (RRMS) in Scotland. This cohort was estimated to capture 45% of persons diagnosed with RRMS in Scotland during the study period^19^ with data collected at baseline and 12 months follow up. Detailed phenotyping of all participants included a wide range of clinical measures and structural brain MRI data. The 3Tesla MRI protocol was performed to obtain T1-weighted, T2-weighted and 2D and 3D FLAIR imaging. Brain tissue volumes and whole brain T2 hyperintense lesion volume (WMH) were derived using longitudinal processing^20^ implemented in FreeSurfer6.0 (http://surfer.nmr.mgh.harvard.edu/) and an in-house adjusted FLAIR thresholding method,^21^ respectively. Full details of the cohort^19^ and the MRI brain acquisition and analysis^22^ have been described elsewhere. Informed consent was obtained prior to study entry according to the Declaration of Helsinki and ethical approval was granted by the South East Scotland Research Ethics Committee 02 (Reference: 15/SS/0233).

### Variable selection

Based on previous literature,^10–16^ network variables were selected to include key clinical, cognitive and imaging metrics previously linked to subjective fatigue in pwMS. Clinical and patient-reported variables were fatigue (Fatigue Severity Scale, FSS),^23^ physical disability (Expanded Disability Status Scale, EDSS),^24^ upper limb dexterity (Nine Hole Peg Test, 9-HPT),^25^ walking speed (Timed 25 Foot Walk, T25FW),^26^ Body Mass Index (BMI), information processing speed (Symbol Digit Modality Test, SDMT),^27^ working memory, auditory information processing and attentional abilities (3 seconds-Paced Auditory Serial Addition Test, PASAT),^28^ anxiety (Generalized Anxiety Disorder-7 instrument, GAD-7),^29^ depression (Patient Heath Questionnaire-9, PHQ-9)^30^ and participant-reported sleep quality measured by an item in Multiple Sclerosis Impact Scale (“SLEEP”).^31^ Imaging variables were expressed as proportions of intracranial volume in native space, including WMH, cortical grey matter volume (“cGM”), basal ganglia volume (“BG”), and thalamus volume (“THALA”). The PHQ-9 has been shown to be a suitable tool to screen for depression with adequate sensitivity and specificity in pwMS.^32^ To control for possible overlap between one PHQ-9 item which assesses feelings of tiredness or reduced energy^30^ and fatigue,^23^ and explore the differential correlations between the remaining PHQ-9 items and fatigue, nine individual PHQ-9 item scores were used in a secondary analysis.^33^ (Table 1) These nine items are identical to the nine core criteria for major depressive disorder in the Diagnostic and Statistical Manual of Mental Disorders, fifth edition (DSM-5).^34^ T25FW, 9-HPT and PASAT were transformed into Z-scores to increase comparability.^19^

**Table 1.**
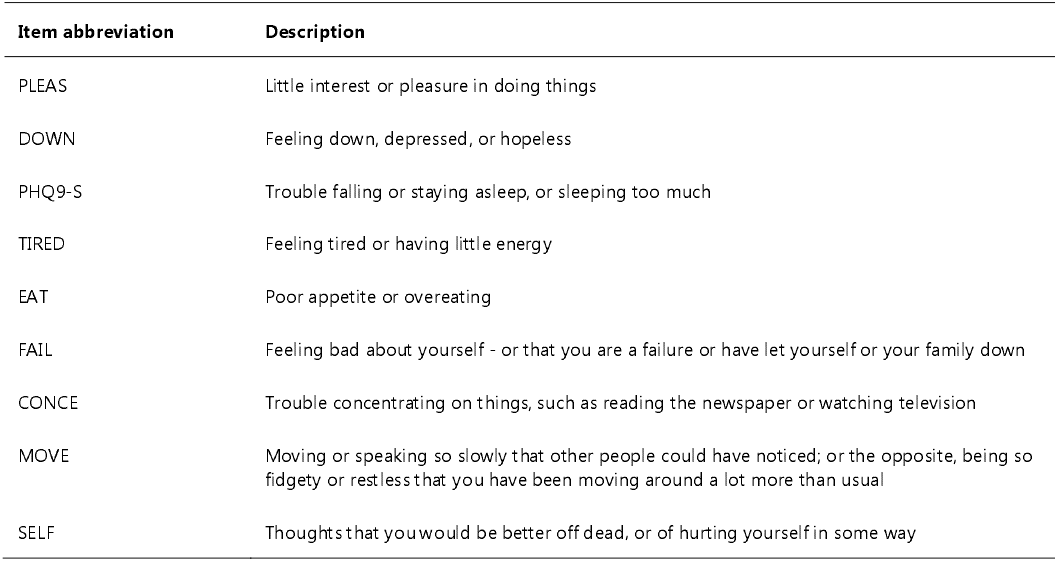
Description of items in the Patient Heath Questionnaire-9 (PHQ-9)

To aid interpretation we classified these variables into five domains as follows: “physical” (EDSS, 9-HPT, T25FW and BMI), “cognitive” (SDMT and PASAT), “affective” (GAD-7 and PHQ-9), “sleep” (SLEEP), “imaging” (WMH, cGM, BG and THALA), and fatigue (FSS). This classification was employed solely to aid interpretation of results and was not included during network estimation.

### Statistical analysis

All analyses were done using R software (v 4.0.2) and R Studio V. 1.2.5042.^35^

Descriptive analysis of demographics and fatigue severity at baseline were conducted. Disease duration was defined as the interval between the patient-described first episode of demyelination, confirmed by clinicians, and the first study visit. Spearman correlations between FSS and the described variables were calculated at study baseline. Bonferroni correction of 13 comparisons was applied in analysis of bivariate Spearman correlations to control family-wise type 1 error rate (α=0.05).

### Network analysis

#### Network estimation

Network analysis aims to identify partial correlations between nodes and represents such correlations as edges. Network estimation was performed via Gaussian graphical models^36^ with Spearman’s correlations. Under this framework, the “Least absolute shrinkage and selection operator”^37^ with the hyperparameter selected by the Extended Bayesian Information Criterion^38^ were applied to prune lower confidence edges from the networks. The tuning parameter was set to 0.5 in all network estimation. This method has been well-documented in previous publications.^39,40^ R package “bootnet”^41^ with the option of EBICglasso was implemented. Visualisation employed the package “qgraph”.^42^ To aid visual comparison, the maximum edge width was set to 0.6 across all networks. Only participants with complete data of all included variables were included in network analysis.

#### Importance of nodes

To quantify the importance of nodes in the network structure, centrality indices, including strength, closeness, betweenness and expected influence were calculated. Strength is the sum of weights connected to a node. As previously defined,^43^ closeness is the reciprocal of the summed length of the shortest paths between a node and all other nodes in the network; betweenness is the number of the shortest paths between two nodes that pass through a node; expected influence is the sum of absolute value of weights connected to a node.

#### Robustness of networks

The robustness of networks was assessed through the methodology proposed by Epskamp et al.^41^ and the R package “bootnet”.^41^ The precision of edge-weight estimates and central indices was obtained by implementing bootstrapping methods with 1000 iterations. The stability of centrality indices, quantified by correlation stability (*CS*) coefficients, was estimated by case-dropping bootstrap. The *CS*-coefficients indicate the maximum percentage of cases that can be dropped to maintain a correlation above 0.7 between subsets’ and original centrality indices.^41^ *CS*-coefficients of 0.5 or higher suggest stable and interpretable results and ranking of centrality indices whereas *CS*-coefficients lower than 0.5 may be interpreted with caution.^41^

#### Post-hoc comparisons and sensitivity analysis

The links between fatigue severity and other variables in the networks were first identified, and then post-hoc analysis via network comparison test (NCT)^44^ was performed to assess whether such links differed between baseline and month 12. NCT is a two-tailed permutation test using a resampling method to construct a distribution under the null hypothesis and test for observed difference. NCT based on 1000 repeats via the R package “NetworkComparisonTest”^44^ was implemented. A Holm-Bonferroni correction was employed to control for multiple testing. Significance level was set at alpha=0.05.

Sensitivity analysis was done by estimating separate networks as described above, stratifying on gender and whether patients had received any Disease Modifying Drugs (DMDs) from baseline to month 12.

## Data availability

Third party data requests will be handled on a case by case basis by the FutureMS steering committee and data custodians as per the approval guidance of the Research Ethics Committee.

## Results

### Baseline characteristics, fatigue severity and bivariate correlations

Among 440 participants enrolled in the FutureMS cohort, all (100%) were included in the analysis of baseline demographics. Participants were newly diagnosed with RRMS (median time from diagnosis to first study visit 60 days, IQR 35 – 96 days) and median disease duration was 1.8 years (IQR 0.9 – 4.6 years). Median age at study inclusion was 36 years (IQR 29 – 45); median EDSS 2 (equivalent to minimal disability) (IQR 1.50 – 3); and 325/440 (73.9%) were female. Geographical and demographic characteristics of the cohort were comparable with those of patients with recently-diagnosed multiple sclerosis in Scotland as a whole, as confirmed by mandatory incidence register for multiple sclerosis in Scotland.^19,45^ Median FSS was 35 (IQR 24 – 47, range 9 – 63), consistent with mild/moderate fatigue severity, and 218 participants (49.5%) reported significant fatigue (FSS ≥ 36).^23^ Fatigue was more severe in females (Female Median 36, IQR 26 – 48; Male Median 30, IQR 19.5 – 44; Wilcoxon rank sum test with continuity correction *p*=0.01), but not significantly correlated with age at study entry (Spearman’s Rho = 0.047, *p*=0.32).

In initial bivariate analyses, FSS significantly correlated with all physical disability, cognitive and affective metrics. In all cases, increasing fatigue was associated with clinically adverse change in the correlated metric (Table 2). Specifically, FSS was strongly correlated with PHQ-9, and moderately correlated with EDSS, GAD-7 and sleep quality. Other correlations were relatively weak.^46^ FSS was not significantly correlated with the included imaging variables.

**Table 2.**
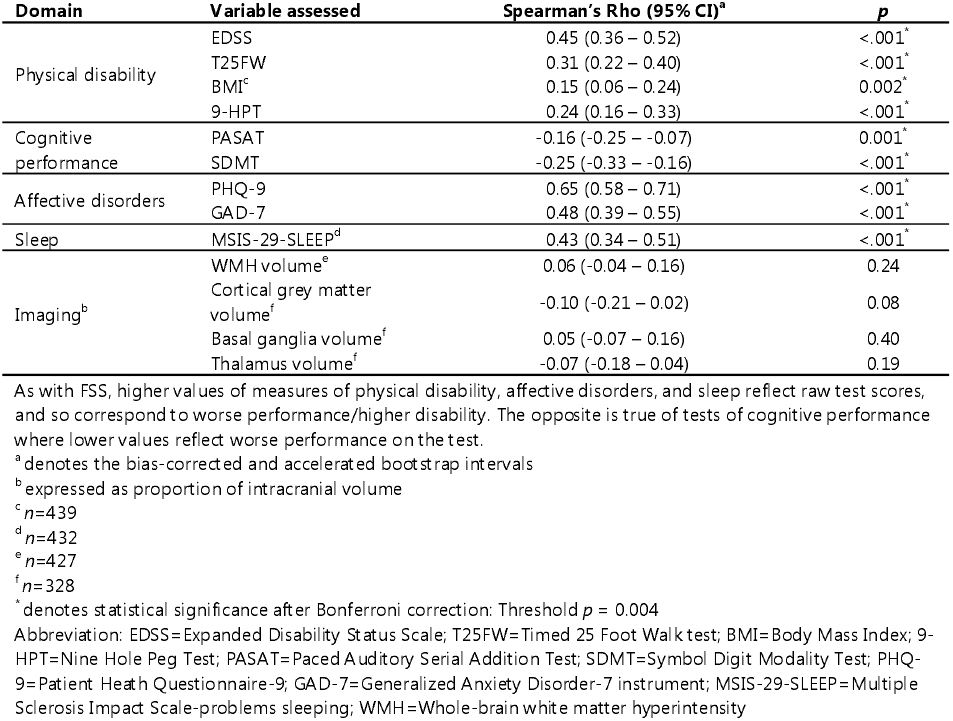
Bivariate correlations of fatigue severity with physical, cognitive, affective, sleep and imaging variables at baseline (n=440)

| Domain | Variable assessed | Spearman's Rho (95% CI)<sup>a</sup> | p |
| --- | --- | --- | --- |
| Physical disability | EDSS | 0.45 (0.36 – 0.52) | <.001<sup>*</sup> |
|  | T25FW | 0.31 (0.22 – 0.40) | <.001<sup>*</sup> |
|  | BMI<sup>c</sup> | 0.15 (0.06 – 0.24) | 0.002<sup>*</sup> |
|  | 9-HPT | 0.24 (0.16 – 0.33) | <.001<sup>*</sup> |
| Cognitive performance | PASAT | -0.16 (-0.25 – -0.07) | 0.001<sup>*</sup> |
|  | SDMT | -0.25 (-0.33 – -0.16) | <.001<sup>*</sup> |
| Affective disorders | PHQ-9 | 0.65 (0.58 – 0.71) | <.001<sup>*</sup> |
|  | GAD-7 | 0.48 (0.39 – 0.55) | <.001<sup>*</sup> |
| Sleep | MSIS-29-SLEEP<sup>d</sup> | 0.43 (0.34 – 0.51) | <.001<sup>*</sup> |
| Imaging<sup>b</sup> | WMH volume<sup>e</sup> | 0.06 (-0.04 – 0.16) | 0.24 |
|  | Cortical grey matter volume<sup>f</sup> | -0.10 (-0.21 – 0.02) | 0.08 |
|  | Basal ganglia volume<sup>f</sup> | 0.05 (-0.07 – 0.16) | 0.40 |
|  | Thalamus volume<sup>f</sup> | -0.07 (-0.18 – 0.04) | 0.19 |

As with FSS, higher values of measures of physical disability, affective disorders, and sleep reflect raw test scores, and so correspond to worse performance/higher disability. The opposite is true of tests of cognitive performance where lower values reflect worse performance on the test.
<sup>a</sup> denotes the bias-corrected and accelerated bootstrap intervals
<sup>b</sup> expressed as proportion of intracranial volume
<sup>c</sup> n=439
<sup>d</sup> n=432
<sup>e</sup> n=427
<sup>f</sup> n=328
<sup>\*</sup> denotes statistical significance after Bonferroni correction; Threshold $p = 0.004$
Abbreviation: EDSS=Expanded Disability Status Scale; T25FW=Timed 25 Foot Walk test; BMI=Body Mass Index; 9-HPT=Nine Hole Peg Test; PASAT=Paced Auditory Serial Addition Test; SDMT=Symbol Digit Modality Test; PHQ-9=Patient Health Questionnaire-9; GAD-7=Generalized Anxiety Disorder-7 instrument; MSIS-29-SLEEP=Multiple Sclerosis Impact Scale-problems sleeping; WMH=Whole-brain white matter hyperintensity

### Baseline and 12-Month Network Analyses

#### Relevant sample characteristics

322 participants (73.2%) at baseline and 323 participants (73.4%) at month 12, had complete data and were therefore included in network analyses. There was no systematic difference in the demographic and clinical measures between participants included in our analysis and the complete cohort (Supplementary Table 1).

The characteristics of this subsample at baseline and month 12 are shown in Table 3. To summarise, we observed a deterioration in EDSS, increase in WMH, and decreases in cGM, BG and Thalamic volume. Nevertheless, improvement was seen in walking speed, upper limb dexterity, and cognitive performance measured by SDMT and PASAT, with reductions in self-reported depression and anxiety. However, there was no significant difference between fatigue severity at baseline and month 12.

**Table 3.** Comparisons of clinical and imaging variables between baseline and month 12.

| Participants eligible for network analysis |  |  |  |
| --- | --- | --- | --- |
|  | Baseline (n = 322) | Month 12 (n = 323) | P<sup>a</sup> |
| FSS | 34 (23.3 – 45.8) | 35 (19 – 50) | 0.556 |
| BMI | 23.30 (26.05 – 30.27) | 23.20 (26.70 – 31.20) | 0.062 |
| EDSS | 2 (1.5 – 3) | 2.5 (2 – 3) | <.001 |
| zT25FW | 0.22 (-0.35 – 0.59) | 0.43 (-0.04 – 0.74) | <.001 |
| z9-HPT | 0.15 (-0.58 – 0.73) | 0.39 (-0.38 – 1.00) | <.001 |
| PHQ-9 | 6 (3 – 11) | 4 (1 – 8.5) | <.001 |
| GAD-7 | 4 (2 – 7) | 3 (1 – 7) | <.001 |
| SDMT | 60.5 (54 – 68) | 62 (54 – 69) | 0.007 |
| zPASAT | 0.32 (-0.16 – 0.73) | 0.53 (-0.06 – 0.87) | <.001 |
| WMH<sup>b</sup> | 0.65 (0.39 – 0.99) | 0.80 (0.59 – 1.20) | <.001 |
| cGM<sup>b</sup> | 37.94 (36.24 – 39.77) | 37.74 (36.16 – 39.47) | <.001 |
| BG<sup>b</sup> | 1.29 (1.20 – 1.40) | 1.26 (1.18 – 1.37) | <.001 |
| THALA<sup>b</sup> | 0.91 (0.83 – 0.97) | 0.90 (0.82 – 0.96) | <.001 |

Data presented above are median (IQR). All Z-scores transformed such that higher values reflect better performance on the test relative to the baseline cohort.
<sup>a</sup> p-values obtained from Wilcoxon signed-rank tests (317 pairs)
<sup>b</sup> expressed as percentage of intracranial volume (%)
Abbreviation: FSS=Fatigue Severity Scale; EDSS=Expanded Disability Status Scale; zT25FW= Z score of Timed 25 Foot Walk test; BMI=Body Mass Index; z9-HPT= Z score of Nine Hole Peg Test; zPASAT=Z score of Paced Auditory Serial Addition Test; SDMT=Symbol Digit Modality Test; PHQ-9=Patient Health Questionnaire-9; GAD-7=Generalized Anxiety Disorder-7 instrument; SLEEP = Multiple Sclerosis Impact Scale-problems sleeping; WMH=Whole-brain white matter hyperintensity volume; cGM=cortical grey matter volume; BG=basal ganglia volume; THALA=thalamus volume

#### Primary networks

Networks at baseline and month 12 (Fig. 1) demonstrated close inter-relationships both within, and between variable groups. With a focus on fatigue in the networks at baseline and month 12, after adjusting for all other variables included in the network, fatigue severity was most strongly connected with PHQ-9 sum score (baseline: 0.34, 95% bootstrap intervals 0.26 – 0.41; month 12: 0.4, 95% bootstrap intervals 0.32 – 0.47) and EDSS (baseline: 0.18, 95% bootstrap intervals 0.08 – 0.27; month 12: 0.14, 95% bootstrap intervals 0.06 – 0.24) (Fig. 1). The strength of these two edges were significantly different from all other edges linked to FSS (Fig S2). Post-hoc analysis examining the links between FSS, PHQ-9 and EDSS did not show significant differences between baseline and month 12 (FSS-PHQ-9: *p*=0.29; FSS-EDSS: *p*=0.57). Weaker connections of FSS were also found with z score of T25FW, GAD-7 and SLEEP (Fig. 1). A weak connection between FSS and PASAT was shown at month 12 only.

**Figure 1.**
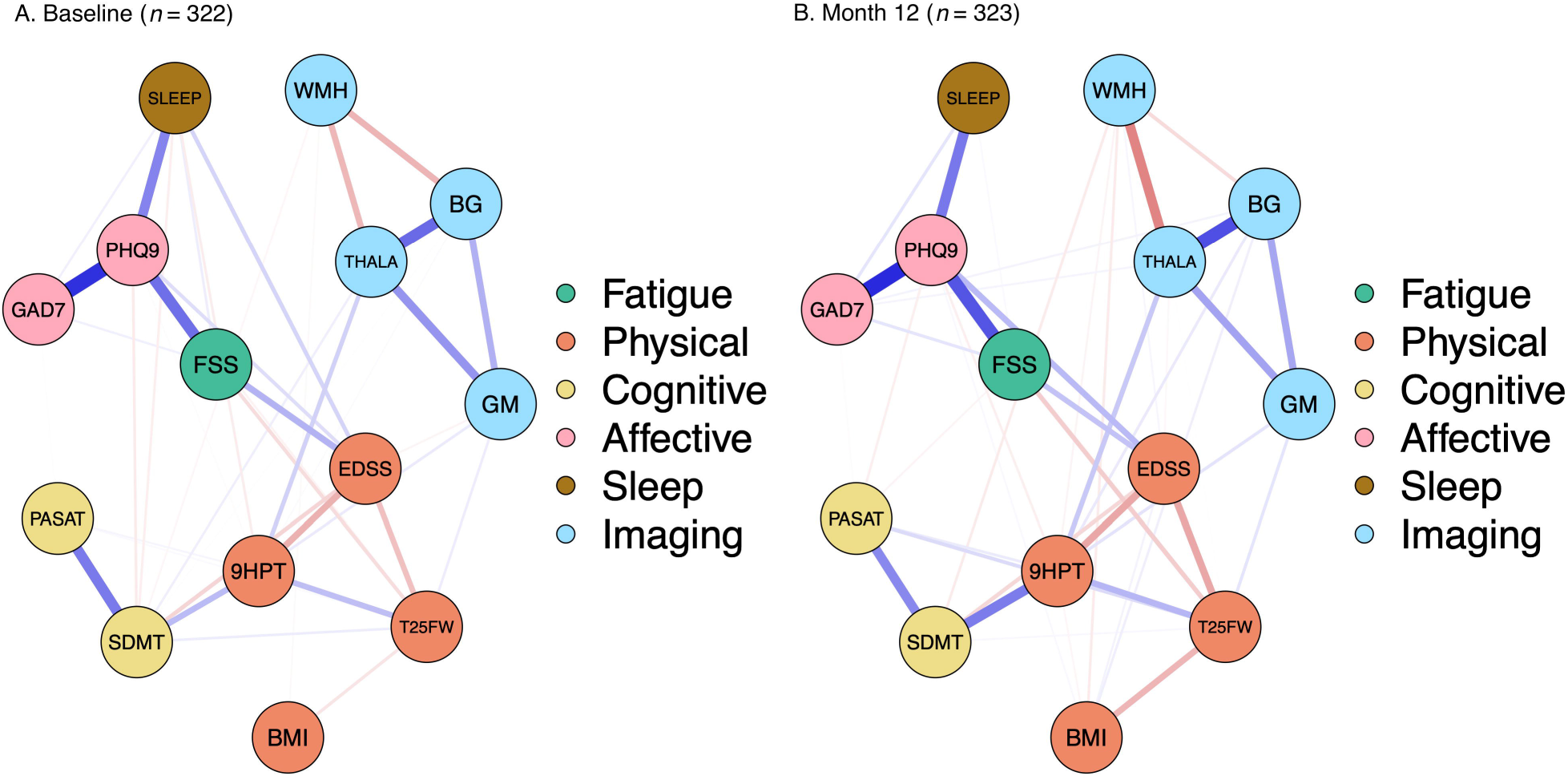
Primary network estimation. Primary networks depicting partial correlations between fatigue and variables of physical disability, affective disorders, cognitive performance, sleep quality, and structural brain imaging. Including PHQ-9 sum scores as measures of depression. (A: Baseline; B: Month 12) Blue edges indicate positive associations and red edges indicate negative associations. The width of the edges is proportional to the absolute value of the edge-weight. The colours of the nodes represent different domains.

#### Secondary networks

In the secondary networks including subscores from individual PHQ-9 items (Fig. 2), fatigue severity was connected to EDSS and to the PHQ-9 item “tiredness”. However, fatigue severity was also specifically connected to PHQ-9 items measuring appetite (“EAT”), subjective concentration deficits (“CONCE”), psychomotor problems (‘MOVE”), and anhedonia (“PLEAS”). Results were consistent at baseline and month 12, with slight differences in very weak associations; for instance, the correlation between FSS and SLEEP was only evident in the secondary network at baseline with PHQ-9 subscores. In contrast, fatigue severity had no links to imaging variables either at baseline or month 12. Complete correlation matrices can be found in supplementary materials (Supplementary Table 2 A-D).

**Figure 2.**
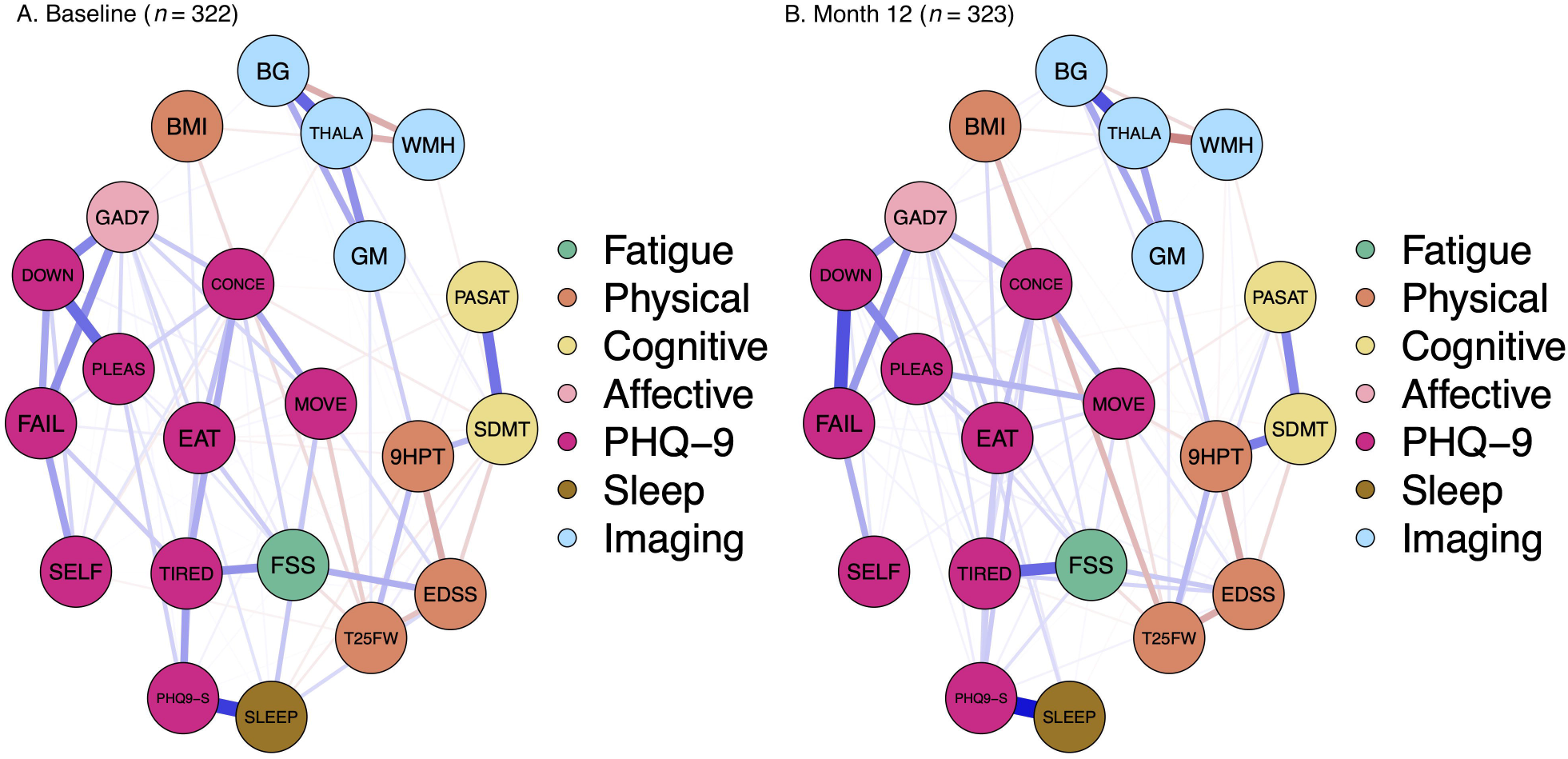
Secondary network estimation. Secondary networks depicting partial correlations between fatigue and variables of physical disability, affective disorders, cognitive performance, sleep quality, and structural brain imaging. Including individual item scores of PHQ-9 as measures of depression. (A: Baseline; B: Month 12) Blue edges indicate positive associations and red edges indicate negative associations. The width of the edges is proportional to the absolute value of the edge-weight. The colors of the nodes represent different domains.

To identify the most central variable in the primary networks at baseline and month 12, only node strength was compared, since *CS*-coefficients of node strength were all above 0.5 across networks. In addition, although there were some negative correlations in the networks, strength and expected influence were highly correlated (*r*=0.7), so only node strength was displayed in the main result. The *CS*-coefficients of all centrality indices are summarized in Supplementary Table 3. In the networks with PHQ-9 sum scores, PHQ-9 had the highest strength (Baseline: 2.24, Month 12: 2.26) (Fig. 3). The differences of node strength between PHQ-9 and other variables were all statistically significant (Supplementary Fig. 3). The node strength of FSS ranked 7^th^ out of 14 both at baseline and month 12 in the networks with PHQ-9 sum scores.

**Figure 3.**
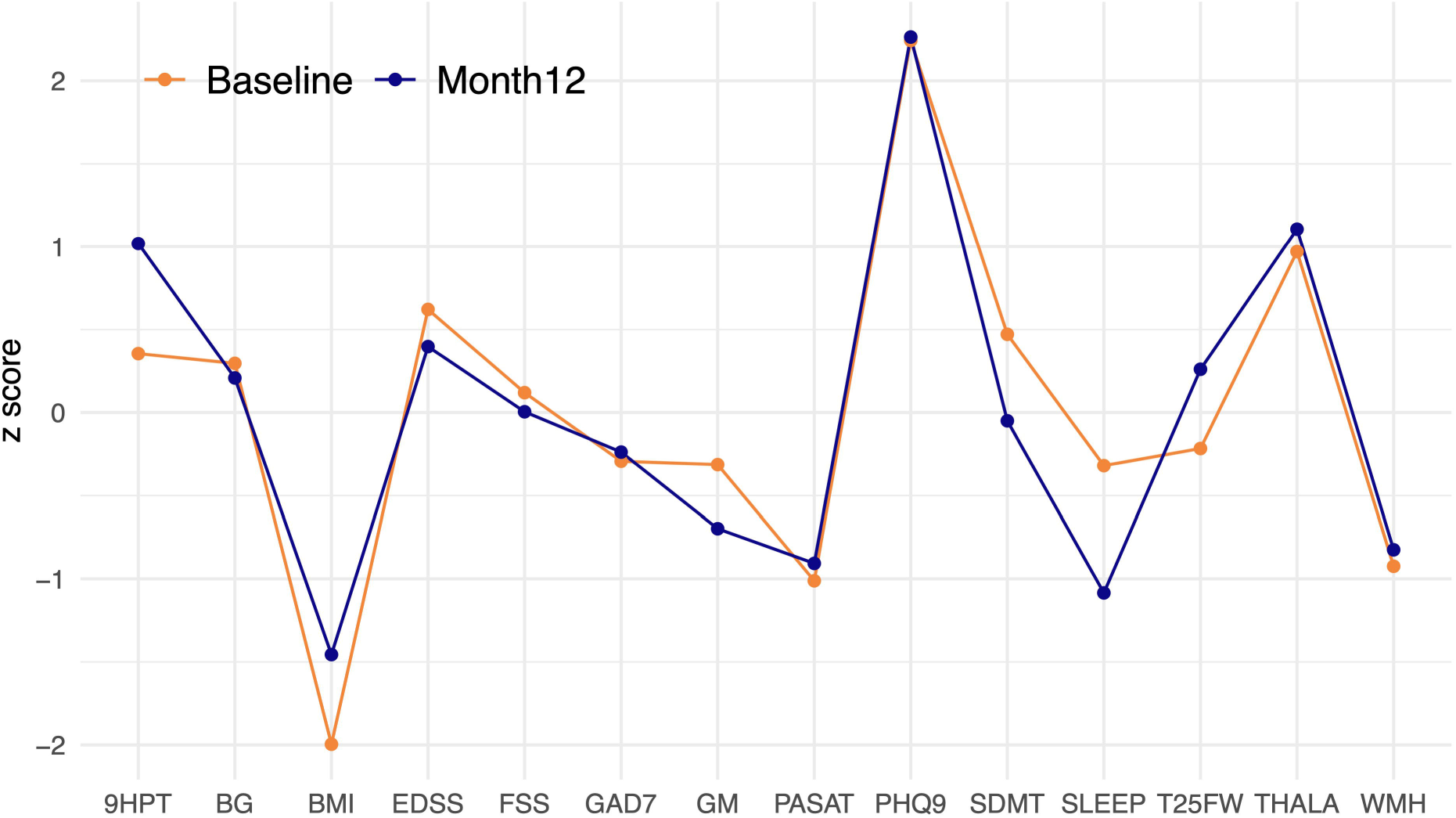
Centrality index – node strength at baseline and month 12. Y-axis displays the z-scores of node strength.

#### Sensitivity analyses

In subgroup networks stratifying on gender (male/female), and on DMD use at follow up (any exposure to DMD at 12-month follow up, yes/no), fatigue severity was associated with EDSS and PHQ-9 in subgroups stratified on DMD initiation, and participant gender, consistent with the cohort as a whole. However, the robustness of networks with male participants (*n*=81) and those who never took DMD (*n*=88) was low (*CS*-coefficients < 0.25) due to small sample sizes. (Supplementary Fig. 6, 7)

## Discussion

In this study, we describe the severity and associations of subjective fatigue in a large representative multi-centre cohort of people recently diagnosed with RRMS. Strikingly, clinically significant fatigue^23^ was present in half of the cohort at baseline. In initial bivariate analyses, we confirmed statistically significant correlations between FSS and a range of physical, cognitive, and affective variables identified in existing literature; though not with T2 lesion volume nor grey matter volumes (of cortex, basal ganglia or thalamus). Use of multivariate network modelling then allowed assessment of the inter-relationships of fatigue with measures of physical disability, cognitive performance, anxiety, depression, subjective sleep quality, whole-brain T2 lesion, cortical grey matter, basal ganglia, and thalamus volume. Network analysis demonstrated that, after accounting for the included variables and their inter-relationships, fatigue severity was most strongly correlated with depression, and with physical disability measured by EDSS, both at baseline and month 12. Several specific depressive symptoms were reproducibly linked to fatigue severity even after statistical correction for an item assessing “tiredness” in the PHQ-9 instrument. Depression had the highest node strength at both baseline and month 12, indicating its strong potential to influence other variables in the networks.

### Links to fatigue – depression, anxiety and physical disability

The link between fatigue and depression has been well-documented. Both are common complaints in pwMS and often coexist.^47^ The association is bidirectional and challenging to disentangle, with identification of fatigue, or depression, increasing the risk of incident depression or fatigue, respectively.^48,49^ Tiredness is one of the nine core symptoms in diagnosing major depressive disorder according to the DSM-5.^34^ In pwMS, a number of mechanistic hypotheses have been proposed to explain the common co-occurrence of fatigue and depression. Firstly, inflammation and cytokine production in response to underlying pathology of multiple sclerosis have been hypothesised to play a role in the experience of both fatigue^50^ and depression^51^ in pwMS, perhaps mediated by effects on brain structures implicated in reward processing.^51–54^ We identified in our cohort a significant relationship between fatigue severity and anhedonia. However, studies of inflammatory biomarkers in serum and CSF have yielded inconsistent results, with no clear biochemical correlate of subjective fatigue in pwMS emerging in studies to date.^8–11^ Secondly, dysregulation of the hypothalamic-pituitary-adrenal axis has been suggested to underlie the coexistence of fatigue and depression in pwMS.^51,53^ A third possible explanation may lie in negatively biased recall, or self-report. Because measurement of both fatigue and depression relies on participant self-report of symptoms, negatively biased cognitive processing^55^ or recall might result in depressed individuals rating their symptoms or functioning more negatively. Such a potential bias might contribute to associations between fatigue and other self-report measures in our study, including anxiety and sleep quality. However, we argue that the subjective experience of fatigue is of importance since it is distinct from objective performance and may reflect metacognitive perception of lack of control over physiological states.^56^ Improved understanding of metacognitive aspects of subjective fatigue may contribute to understanding the promising results of cognitively-focussed therapies in treatment of multiple sclerosis-related fatigue.^57^

Apart from depression sum score, subjective fatigue may be differentially linked to specific depressive symptoms.^58^ Among nine core depressive symptoms,^30,34^ and after controlling for an item measuring tiredness, we found four of these (anhedonia, subjective concentration deficits, subjectively altered speed of movement, and appetite) which may be specifically related to subjective fatigue. These findings were consistent with previous literature which suggested that separate depressive symptoms are distinct in biological mechanisms, risk factors and impacts on impairment.^58^ Future studies are needed to characterise the relationships between fatigue and specific depressive symptoms in detail.

The link between fatigue and EDSS may also be bidirectional. It has been hypothesised both that fatigue may occur as a consequence of disease severity,^59^ and that, conversely, fatigue-related fear and avoidance might lead to increased disease severity and physical disability.^60^ A large retrospective cohort study^61^ found that fatigue at study enrolment was a significant predictor of later sustained EDSS worsening. However, such a link has not been confirmed by other literature.^10,11^ Heterogeneity of previous findings may be linked to sample size and publication bias, as well as variation in fatigue measures employed, disease duration, and average EDSS scores.^10^ Our study suggests that subjective fatigue remains correlated with physical disability as measured by EDSS, even after adjustment for a number of other variables, and that this finding remains robust over the course of one year, in the early disease course of RRMS.

### Absent links to fatigue in networks – cognitive performance and brain imaging

Absent links in the estimated networks could also yield insights in understanding multiple sclerosis-related fatigue. The discrepancy between the results of bivariate correlations and network analyses could have several possible explanations. First, the network models in our study imposed a sparse network structure. It is thus possible that some weak links were eliminated from the network estimation. Secondly, this may reflect that some apparent links in bivariate correlations are in fact mediated by other variables. For instance, the bivariate correlation between fatigue and BMI may be accounted for by common links with walking speed.^62^ Furthermore, the absence of correlations between fatigue severity and objective cognitive performance in our estimated networks may have further implications. For example, in the secondary networks with PHQ-9 subscores (which included an item assessing subjective difficulty in concentrating), fatigue severity was not linked to objective cognitive performance measured by SDMT or PASAT, whilst it was connected with subjective difficulty in concentrating. These findings suggest, in our cohort, that subjective cognitive difficulty and objective cognitive performance may be dissociated, with subjective but not objective deficits linked to fatigue severity. Comparable dissociation between objective and subjective estimates of cognitive performance has previously been described in association with fatigue in pwMS.^14^ Cognitive performance is one aspect of fatigability^2,53^ and therefore this observation further adds to understanding of links between subjective fatigue, and fatigability.^56^

Furthermore, our study did not find an association between fatigue severity and structural brain imaging variables, using either bivariate or network analysis. A recent review^54^ described that the associations between fatigue and global brain structures, including white matter lesion volume and grey matter volume, were not significant, or lost significance after controlling for important confounding variables, such as EDSS.^11^ The absence of a relationship between fatigue severity, and volumes of basal ganglia and thalamus in our findings was however different from some previous studies.^54^ A study examined patients with RRMS and showed significant atrophy of basal ganglia and thalamus in fatigued compared to non-fatigued participants.^63^ Moreover, this study showed that the cognitive and physical domains of fatigue were both correlated with striatal volume, and with cortical thickness of different brain areas, including the parietal and frontal lobes.^63^ Furthermore, two reviews summarised brain imaging findings in multiple sclerosis-related fatigue and similarly concluded that macro/microstructural and functional changes involving the cortico-striato-thalamo-cortical loop might contribute to the pathogenesis of fatigue in pwMS.^16,54^ Reasons for contrasting results in our study, in comparison to previous publications, could include methodological considerations such as the disease duration and disease course of enrolled patients.^11,63,64^ Furthermore, brain structural changes related to fatigue in early RRMS might only be apparent at a microstructural or functional level rather than at the macrostructural levels examined in our study.

### Limitations and strengths

There are some limitations in our study. First, in this study we measured fatigue severity using FSS, which despite validation for use in pwMS, has been suggested to preferentially focus on physical aspects of fatigue.^65^ However, we note a strong association between fatigue severity as described by FSS, and other correlates of fatigue, particularly depressive symptomatology, in the current study. Secondly, causality cannot be inferred from cross-sectional analyses included in this study. Future studies including data collection at multiple time points to build longitudinal networks are needed. Lastly, due to the availability of data in this cohort, we could not include all variables that could possibly link to fatigue. For instance, pain has been found to be closely linked to both fatigue and depression.^52^ Some serum and CSF biomarkers (such as IL-6),^8–11^ as discussed above, might also relate to fatigue. Future studies that include measures of pain and serum/CSF biomarkers might further mechanistic understanding.

Nevertheless, this study has four major strengths. First, the data analysed in our study were drawn from the FutureMS cohort study, which showed geographically and demographically representative coverage of Scottish patients diagnosed with RRMS.^19^ Secondly, none of the study participants had received DMDs at study baseline, which allowed us to examine the associations of interest without effects from DMDs at study baseline, and to contrast findings with those one year later, when 72.7% had commenced disease modifying therapy. Thirdly, in light of the deep phenotyping in the FutureMS cohort, we were able to employ a wide range of relevant clinical, neuropsychological and imaging phenotyping which allows a unique opportunity to examine the associations of fatigue with a broad range of relevant variables. Lastly, use of a network approach allows for assessing the dynamic relationships between fatigue, clinical measures, and brain structure assessed by MRI, after correction for all other variables in the network. This method is well grounded in psychological research, but relatively new in the area of neurological diseases.

### Implications for future studies and clinical practice

From a clinical perspective, the strong links identified between subjective fatigue and depression, particularly specific depressive symptoms (anhedonia, subjective concentration deficits, subjectively altered speed of movement, and appetite), may assist focussed recognition and assessment of comorbid fatigue and depression, and suggest that depressive symptoms should be rigorously assessed in pwMS reporting fatigue. Besides the beneficial outcomes that have been shown in interventions simultaneously targeting fatigue and depression,^66^ a recent study found that early intervention in reducing depressive symptoms was associated with overall reduction in fatigue severity, and vice versa.^67^ The findings in our study also suggested that a focus on identification and modification, where possible, of depression and physical disability may be particularly important in clinical treatment of subjective fatigue. Moreover, only weak or absent evidence of relationships between subjective fatigue and sleep quality, cognitive performance, or structural brain imaging variables were found, which could also provide valuable insights on pathogenesis. Furthermore, post-hoc analysis showed that the links between fatigue severity and key depressive symptoms were not significantly different between baseline and month 12, suggesting that the underlying contributory factors of subjective fatigue remained stable in this timeframe. However, longer-duration longitudinal studies are needed to determine whether these key depressive symptoms are specifically linked to subjective fatigue in pwMS, and whether there is a shift in correlations of fatigue severity throughout the disease course. From a research perspective, data-driven multivariate approaches such as network analysis could be especially useful in guiding selection of the most important research and clinical targets in multiple sclerosis-related fatigue, particularly where relevant variables are known to be inter-related. The stability of estimated networks can furthermore usefully be quantified, along with strength and influence of network nodes.

### Conclusion

In summary, we demonstrate the considerable burden of subjective fatigue for people with RRMS even early in the disease course. We also describe consistent correlations between subjective fatigue severity and depressive symptoms, as well as with physical disability measured by EDSS in early RRMS. In contrast, network analyses did not support significant associations between fatigue severity and objective cognitive performance or structural brain imaging variables. Subjective fatigue appears to be preferentially linked to specific key depressive symptoms in our cohort. The findings in this study provide additional evidence in understanding the aetiology of fatigue in multiple sclerosis, and underline the importance of fatigue and its association with specific depressive symptoms in both a clinical and research context.

## Supporting information

STROBE

## Acknowledgements

This study is indebted to the FutureMS participants. We would like to thank non-author contributors of the FutureMS Consortium and clinical collaborators within neurology departments across NHS Scotland. We would also like to acknowledge Ms Nicole White’s contribution to the brain imaging used in this study. With thanks to FutureMS, hosted by Precision Medicine Scotland Innovation Centre (PMS IC) and funded by a grant from the Scottish Funding Council to PMS IC and Biogen Idec Ltd Insurance.

## Abbreviations

9-HPT: Nine Hole Peg Test
BG: basal ganglia volume
BMI: Body Mass Index
CI: confidence interval
DMDs: Disease Modifying Drugs
DSM-5: Diagnostic and Statistical Manual of Mental Disorders, fifth edition
EDSS: Expanded Disability Status Scale
FSS: Fatigue Severity Scale
GAD7: Generalized Anxiety Disorder-7
cGM: cortical grey matter volume
PASAT: Paced Auditory Serial Addition Test
PHQ-9: Patient Heath Questionnaire-9
pwMS: people with MS
RRMS: relapsing-remitting multiple sclerosis
SDMT: Symbol Digit Modality Test
SLEEP: Multiple Sclerosis Impact Scale-problems sleeping
T25FW: Timed 25 Foot Walk test
THALA: thalamus volume
WMH: Whole-brain white matter hyperintensity volume

