## Supplemental Data for "Data-driven analysis shows robust links between fatigue and depression in early multiple sclerosis"

### Supplementary Materials

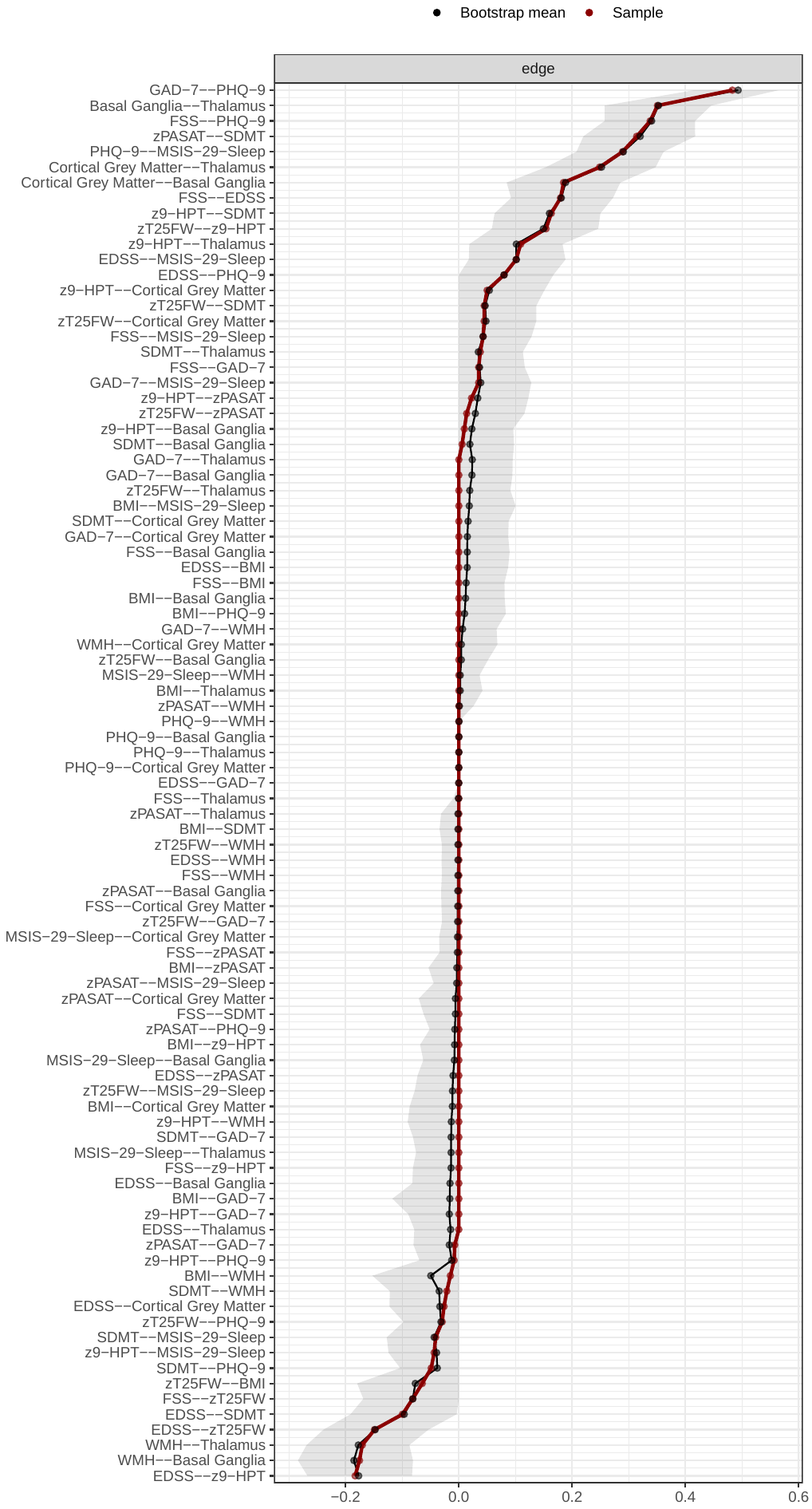

**Supplementary Fig. 1A Summaries of bootstrapped sampling distributions separately for the weight of each edge at baseline with PHQ-9 sum scores.** The red dots indicate the weight of each edge in this study population. The black dots are plotted at the arithmetic means of the boot strapped sampling distributions. The grey shading indicates the 0.025 and 0.975 quantiles of the bootstrapped sampling distribution.

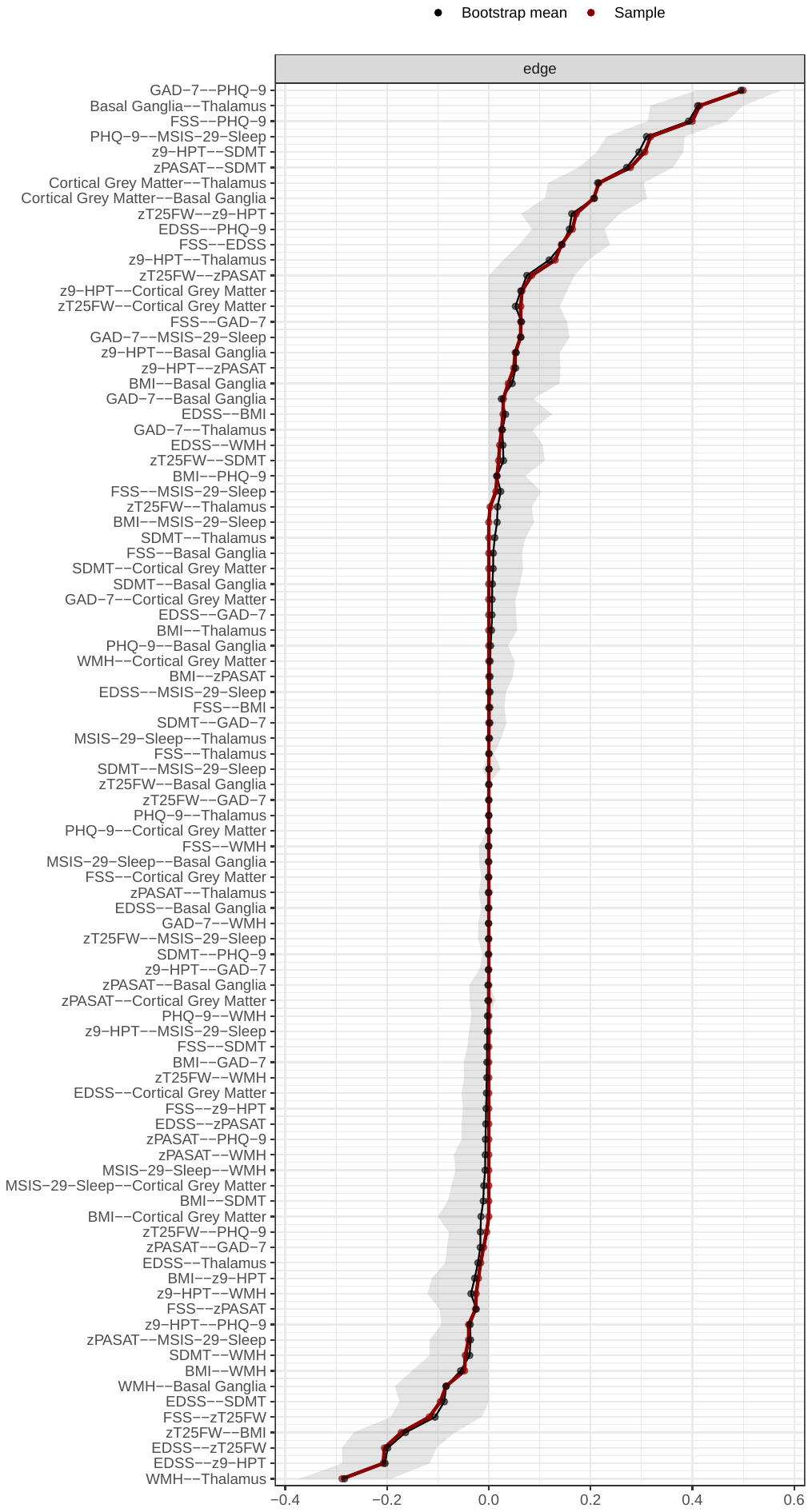

**Supplementary Fig. 1B Summaries of bootstrapped sampling distributions separately for the weight of each edge at month 12 with PHQ-9 sum scores**. The red dots indicate the weight of each edge in this study population. The black dots are plotted at the arithmetic means of the boot strapped sampling distributions. The grey shading indicates the 0.025 and 0.975 quantiles of the bootstrapped sampling distribution.

| 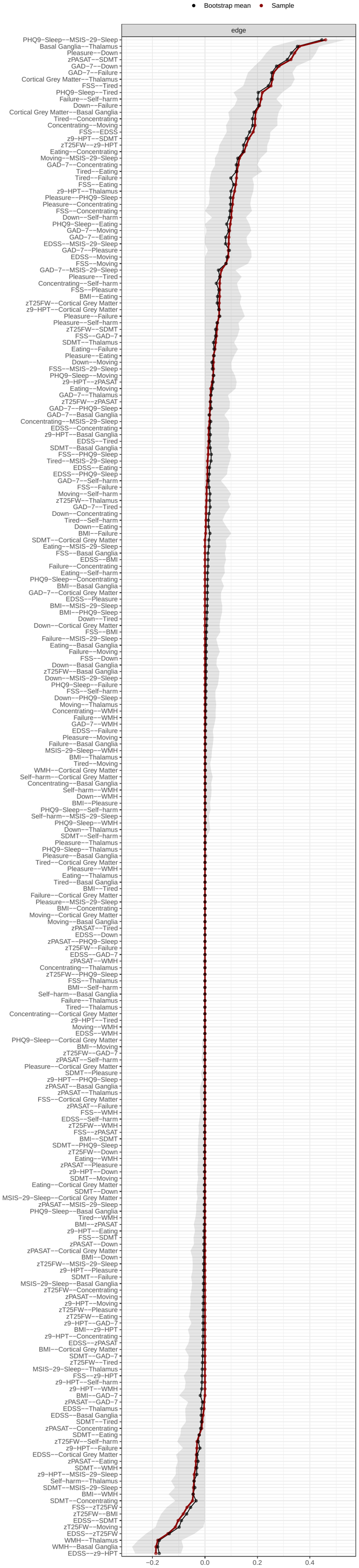 | **Supplementary Fig. 1C Summaries of bootstrapped sampling distributions separately for the weight of each edge at baseline with PHQ-9 subscores.** The red dots indicate the weight of each edge in this study population. The black dots are plotted at the arithmetic means of the boot strapped sampling distributions. The grey shading indicates the 0.025 and 0.975 quantiles of the bootstrapped sampling distribution. |
| --- | --- |

| 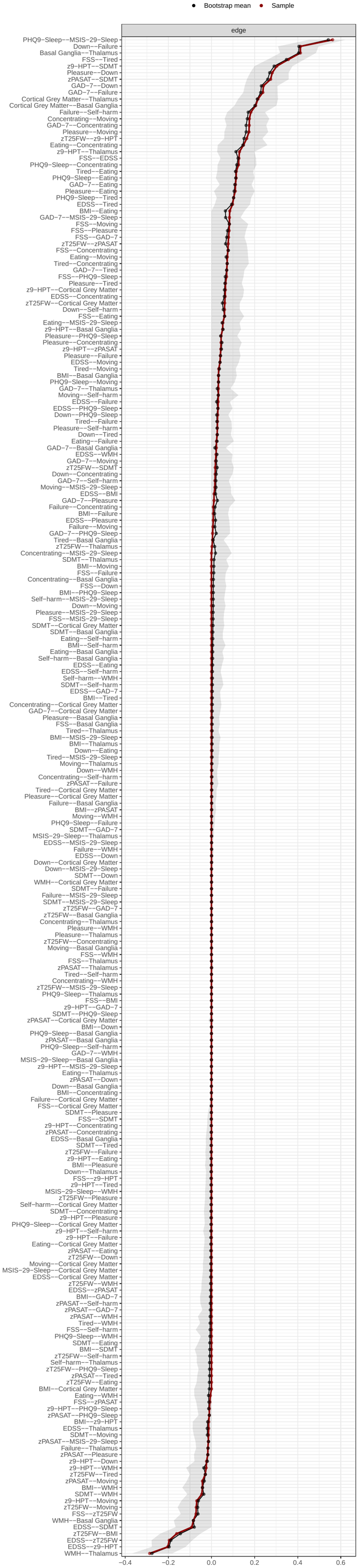 | **Supplementary Fig. 1D Summaries of bootstrapped sampling distributions separately for the weight of each edge at month 12 with PHQ-9 subscores.** The red dots indicate the weight of each edge in this study population. The black dots are plotted at the arithmetic means of the boot strapped sampling distributions. The grey shading indicates the 0.025 and 0.975 quantiles of the bootstrapped sampling distribution. |
| --- | --- |

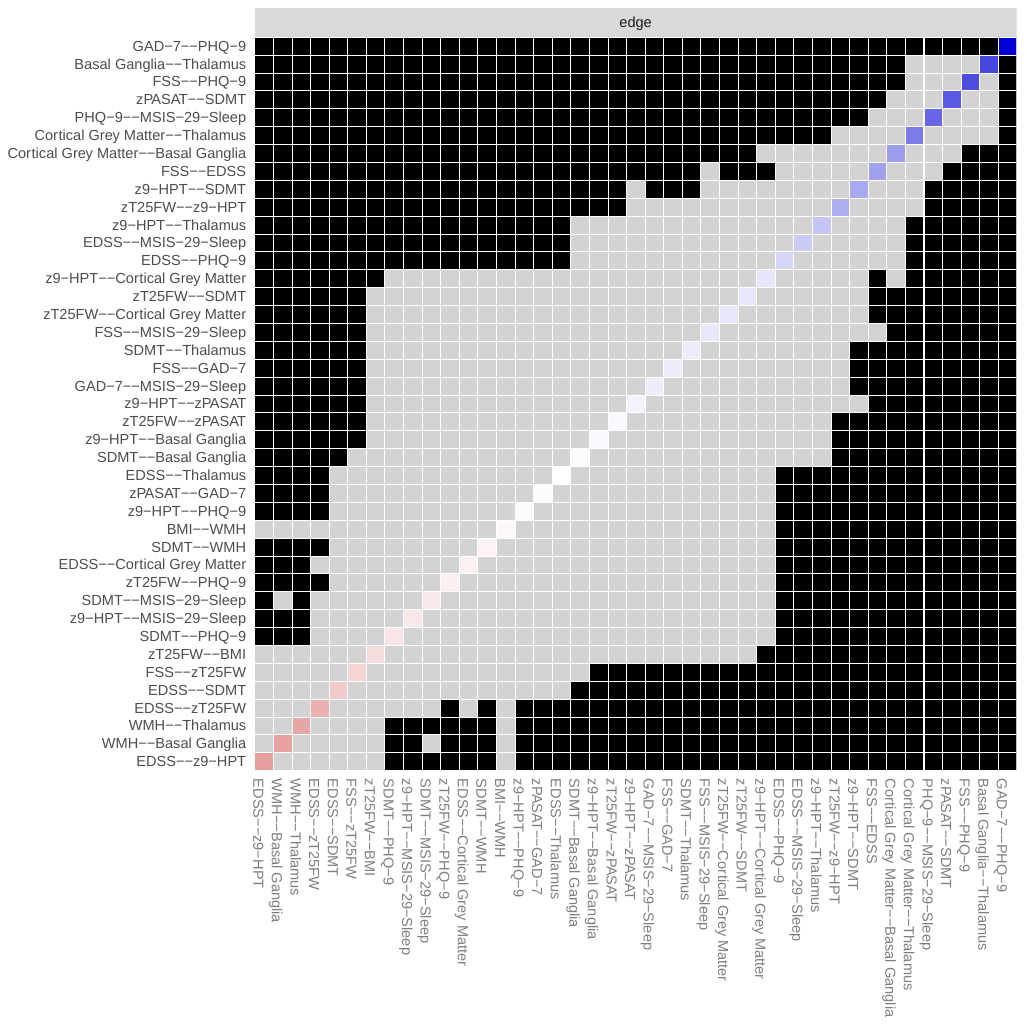

**Supplementary Fig. 2A Bootstrapped tests for difference between non-zero edge-weights in the estimated network at baseline with PHQ-9 sum scores (α = 0.05).** Each box in the figure above is the test result between corresponding two edges at x-axis and y-axis. Black box indicates the two edge-weights are significantly different, while gray box indicates the two edge-weights are not significantly different. Coloured boxes correspond to the colour of the edges in Fig. 1 & 2, in which blue edges indicate positive correlations and red edges indicate negative correlations.

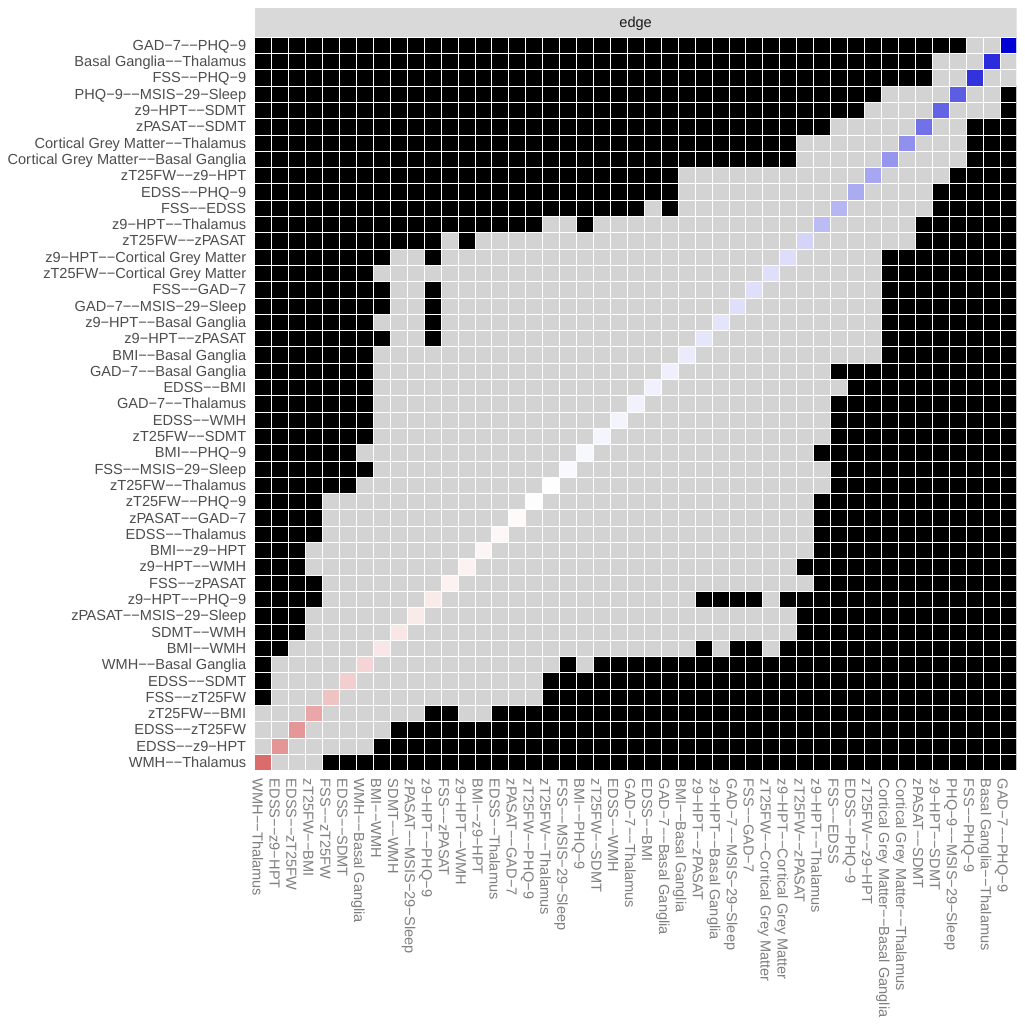

**Supplementary Fig. 2B. Bootstrapped tests for difference between non-zero edge-weights in the estimated network at month 12 with PHQ-9 sum scores (α = 0.05).** Each box in the figure above is the test result between corresponding two edges at x-axis and y-axis. Black box indicates the two edge-weights are significantly different, while gray box indicates the two edge-weights are not significantly different. Coloured boxes correspond to the colour of the edges in Fig. 1 & 2, in which blue edges indicate positive correlations and red edges indicate negative correlations.

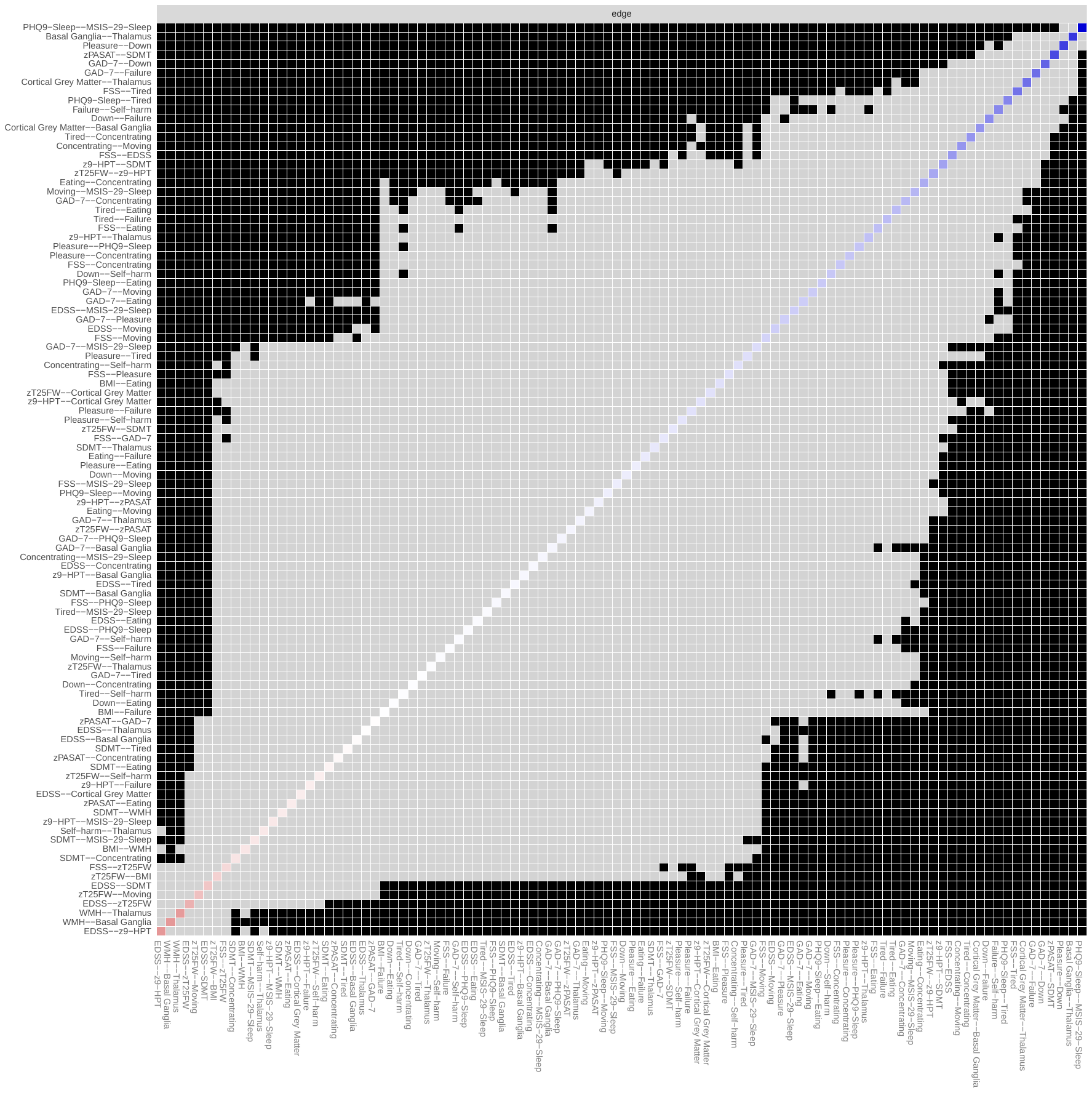

**Supplementary Fig. 2C Bootstrapped tests for difference between non-zero edge-weights in the estimated network at baseline with PHQ-9 subscores (α = 0.05).** Each box in the figure above is the test result between corresponding two edges at x-axis and y-axis. Black box indicates the two edge-weights are significantly different, while gray box indicates the two edge-weights are not significantly different. Coloured boxes correspond to the colour of the edges in Fig. 1 & 2, in which blue edges indicate positive correlations and red edges indicate negative correlations.

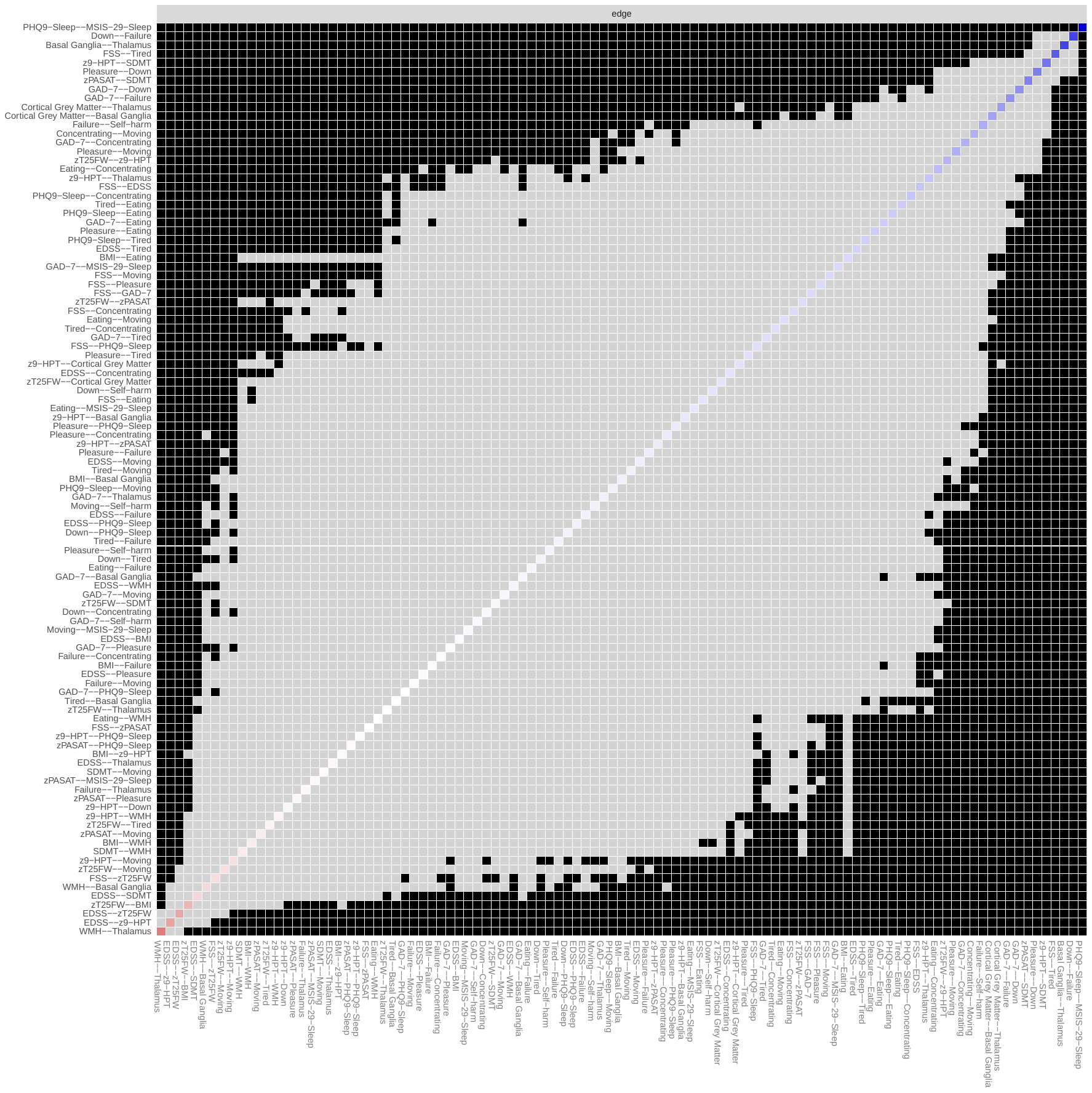

**Supplementary Fig. 2D Bootstrapped tests for difference between non-zero edge-weights in the estimated network at month 12 with PHQ-9 subscores (α = 0.05).** Each box in the figure above is the test result between corresponding two edges at x-axis and y-axis. Black box indicates the two edge-weights are significantly different, while gray box indicates the two edge-weights are not significantly different. Coloured boxes correspond to the colour of the edges in Fig. 1 & 2, in which blue edges indicate positive correlations and red edges indicate negative correlations.

| A. Baseline  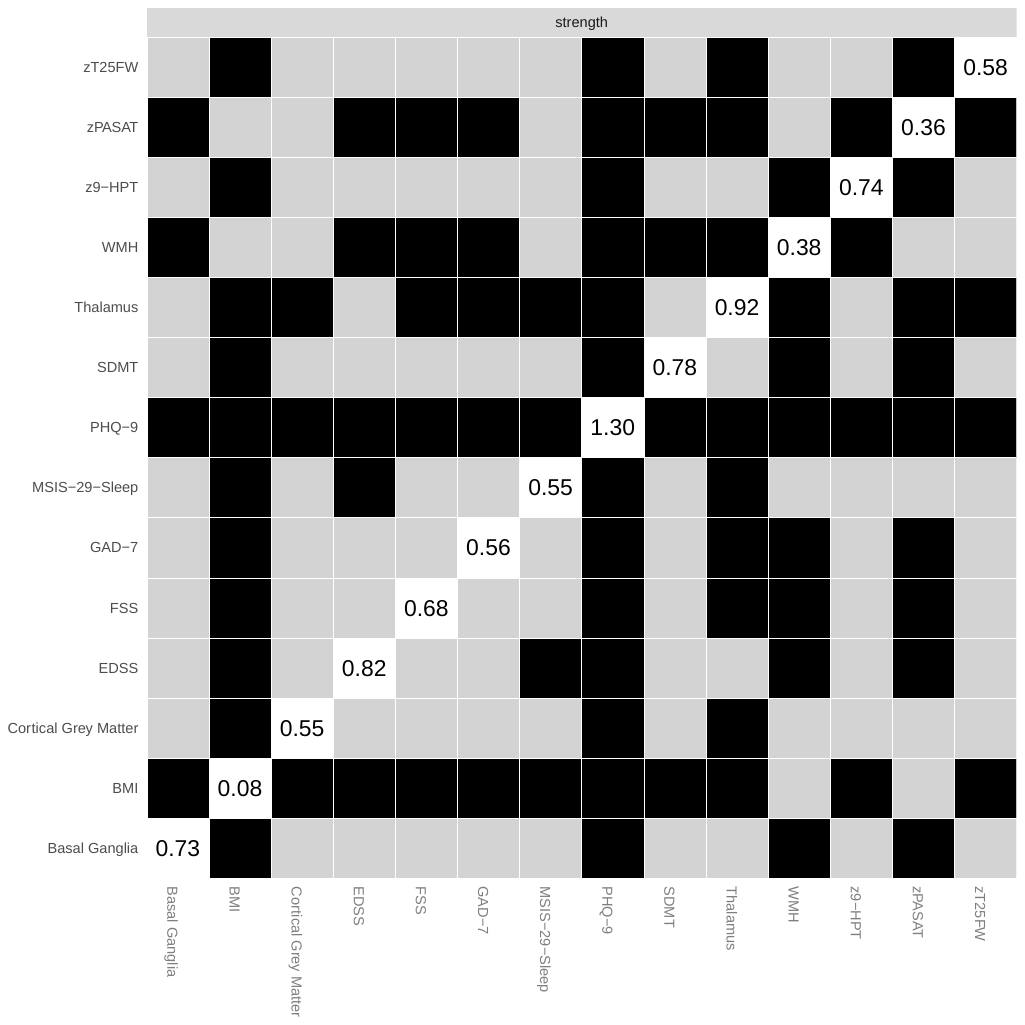 | B. Month 12  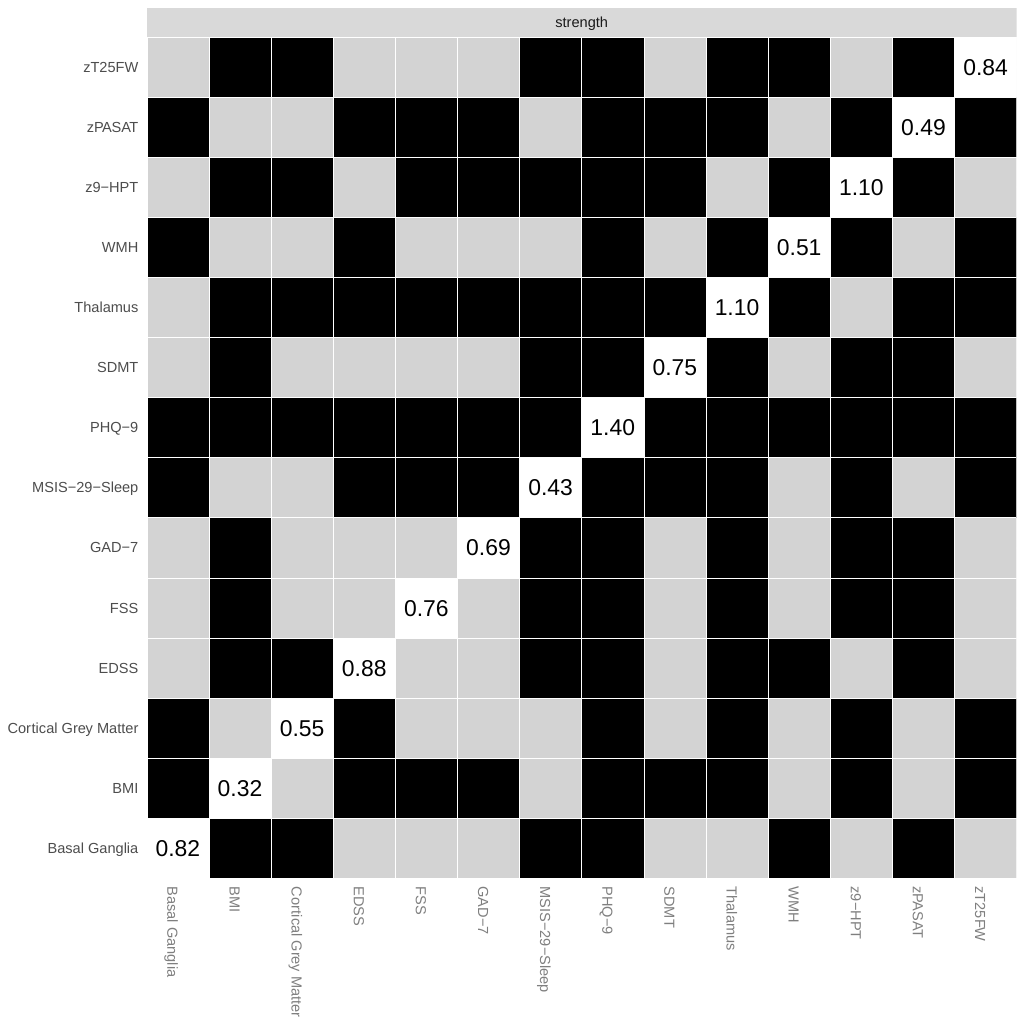 |
| --- | --- |
| C. Baseline, PHQ-9 subscores  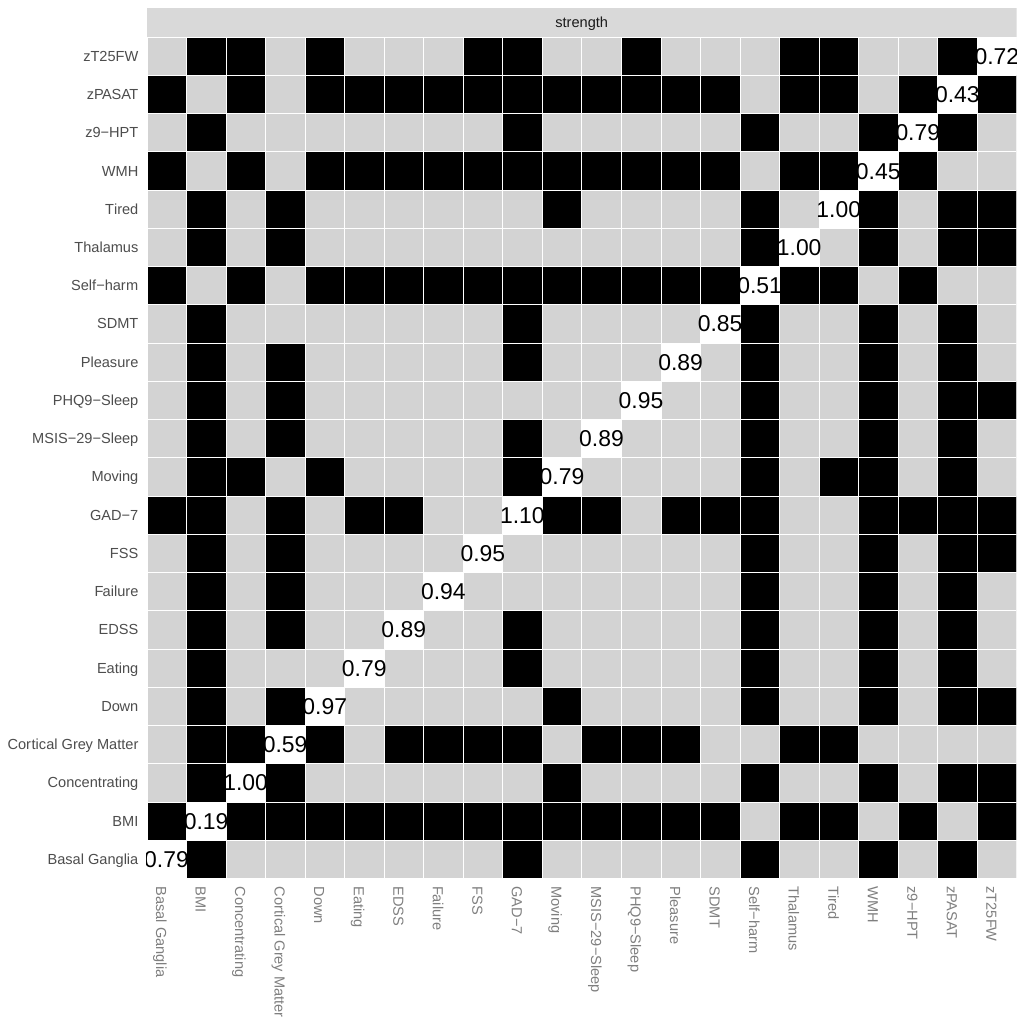 | D. Month 12, PHQ-9 subscores  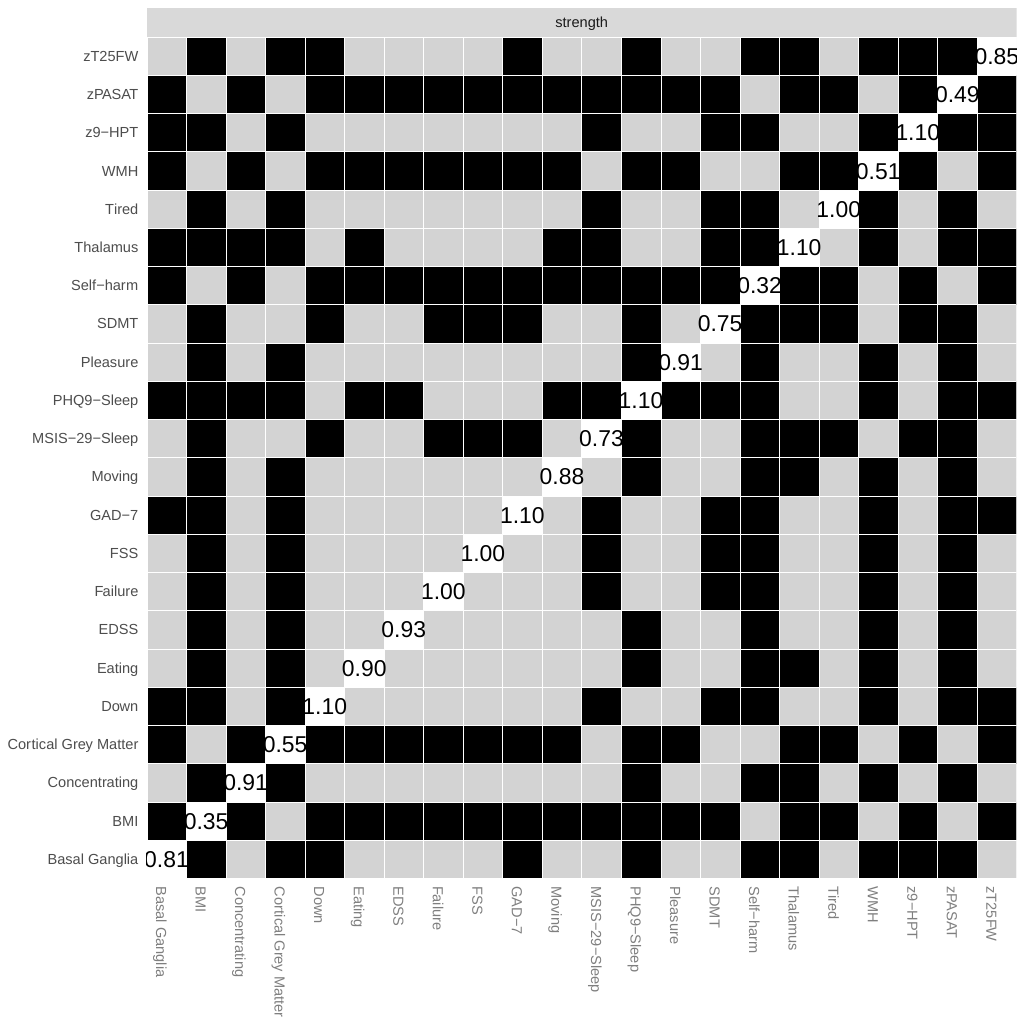 |

**Supplementary Fig. 3 Bootstrapped tests for difference between node strength of the variables in the estimated networks** (A: Baseline with PHQ-9 sum scores; B: Month 12 with PHQ-9 sum scores; C: Baseline with PHQ-9 subscores; D: Month 12 with PHQ-9 subscores) (α = 0.05). Each box in the figure above is the test result between corresponding two nodes at x-axis and y-axis. Black box indicates the two nodes are significantly different in strength, while gray box indicates the two nodes are not significantly different in strength. White boxes in the diagonal lines show the value of node strength.

| A  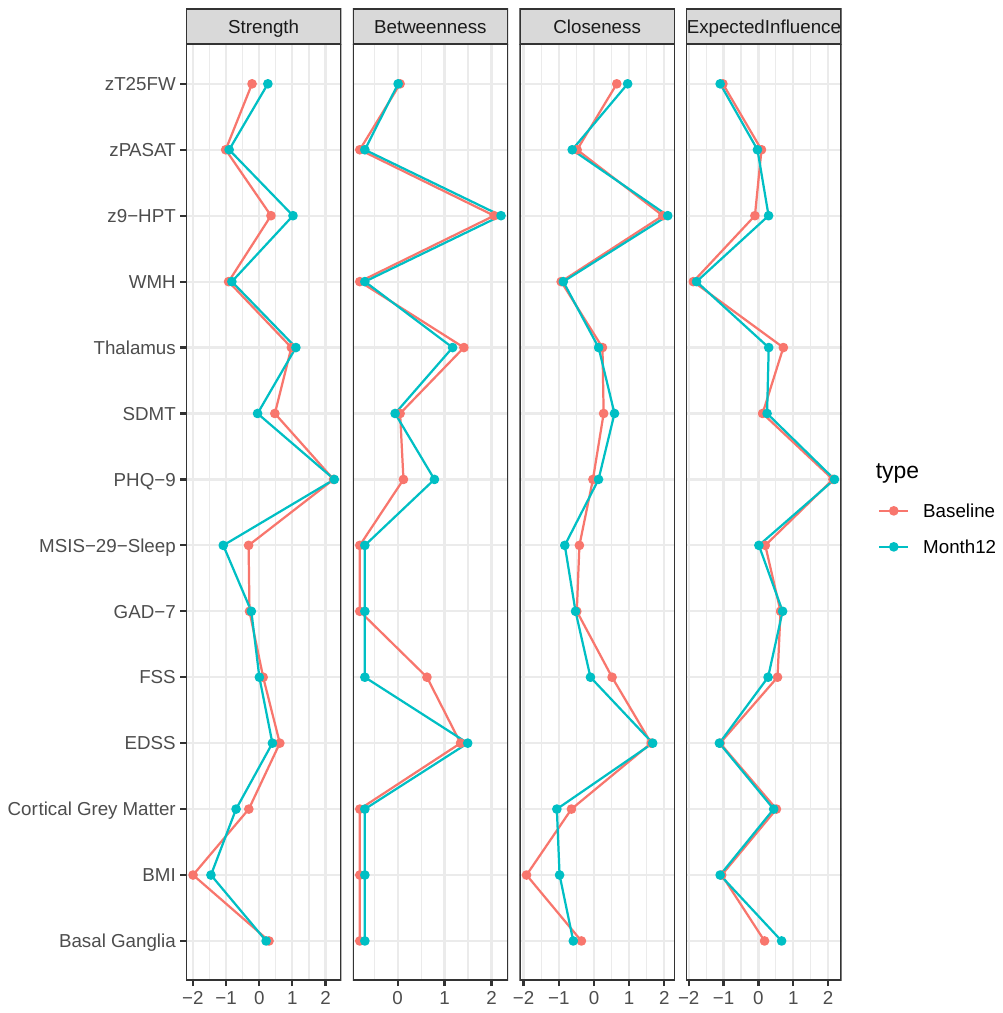 | **Supplementary Fig. 4 Centrality indices at baseline and month 12** (A: PHQ-9 sum scores; B: PHQ-9 subscores). X-axis represent the z-scores of centrality indices. Abbreviation: FSS=Fatigue Severity Scale; EDSS=Expanded Disability Status Scale; zT25FW= Z Scores of Timed 25 Foot Walk test; BMI=Body Mass Index; z9-HPT=Z Scores of Nine Hole Peg Test; zPASAT= Z Scores of Paced Auditory Serial Addition Test; SDMT=Symbol Digit Modality Test; GAD7=Generalized Anxiety Disorder-7 instrument; PHQ-9=Patient Heath Questionnaire-9: PLEAS= little interest or pleasure in doing things, DOWN= feeling down, depressed, or hopeless, PHQ9-S= trouble falling or staying asleep, or sleeping too much, TIRED= feeling tired or having little energy, EAT= poor appetite or overeating, FAIL= feeling bad about yourself - or that you are a failure or have let yourself or your family down, CONCE= trouble concentrating on things, such as reading the newspaper or watching television, MOVE= moving or speaking so slowly that other people could have noticed; or the opposite, being so fidgety or restless that you have been moving around a lot more than usual, SELF= thoughts that you would be better off dead, or of hurting yourself in some way; SLEEP = Multiple Sclerosis Impact Scale-Problems sleeping; WMH=Whole-brain white matter hyperintensity volume; cGM=cortical grey matter volume; BG=basal ganglia volume; THALA=thalamus volume |
| --- | --- |
| B  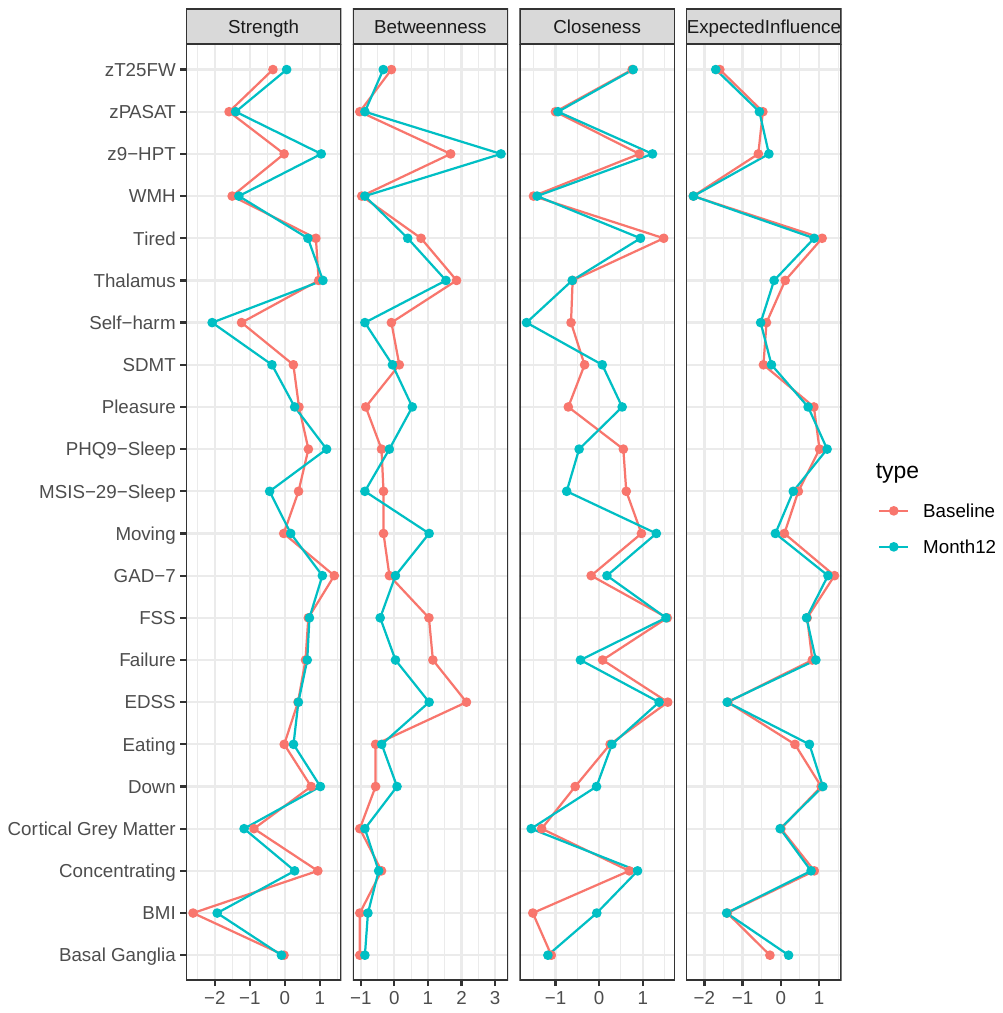 |  |

| A. Baseline  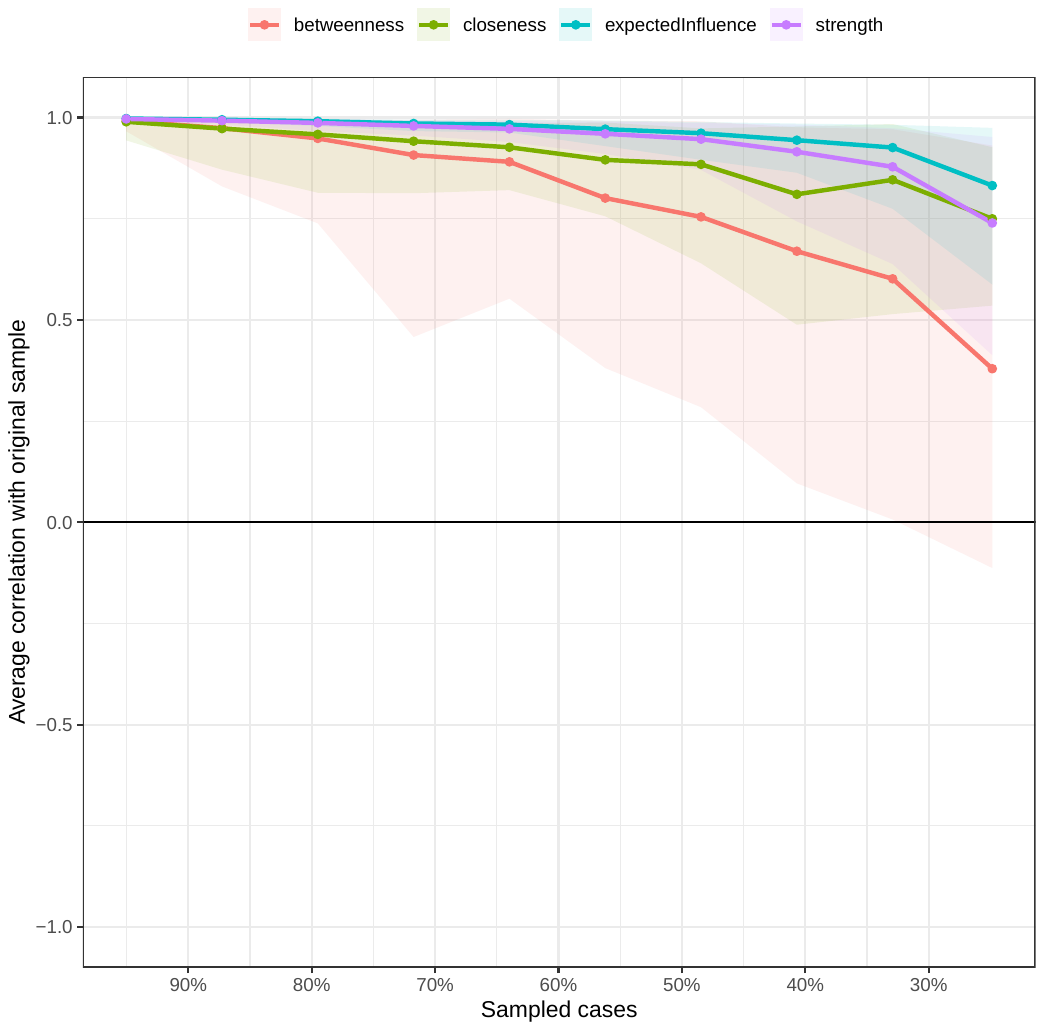 | B. Month 12  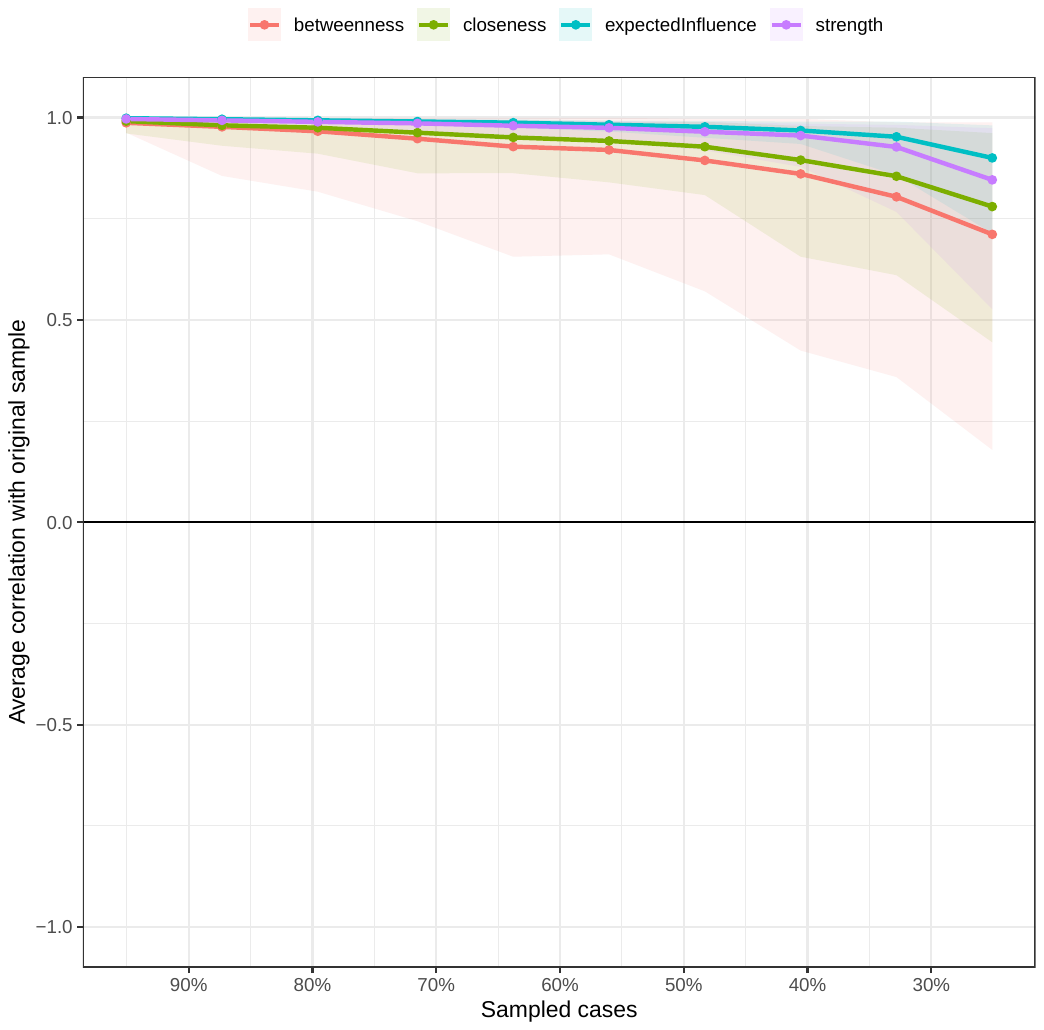 |
| --- | --- |
| C. Baseline, PHQ-9 subscores  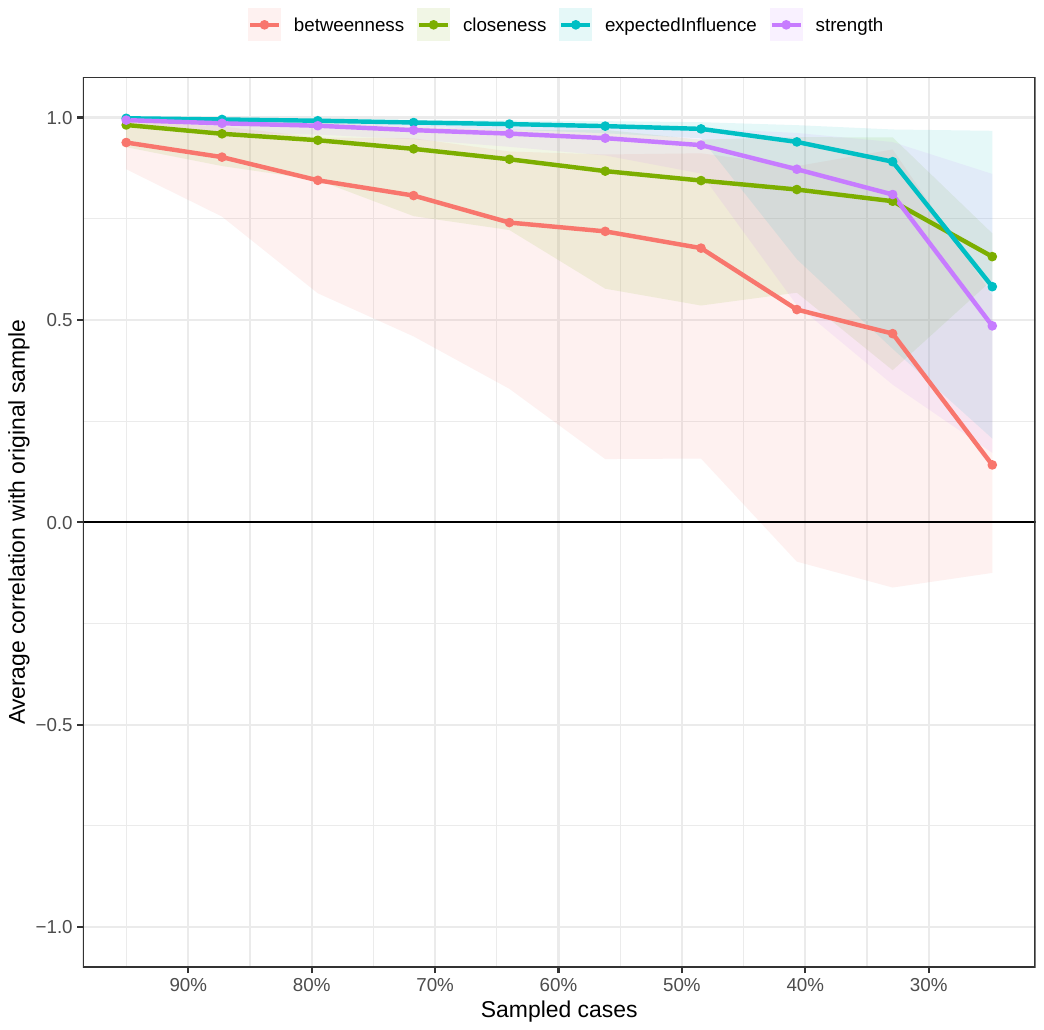 | D. Month 12, PHQ-9 subscores  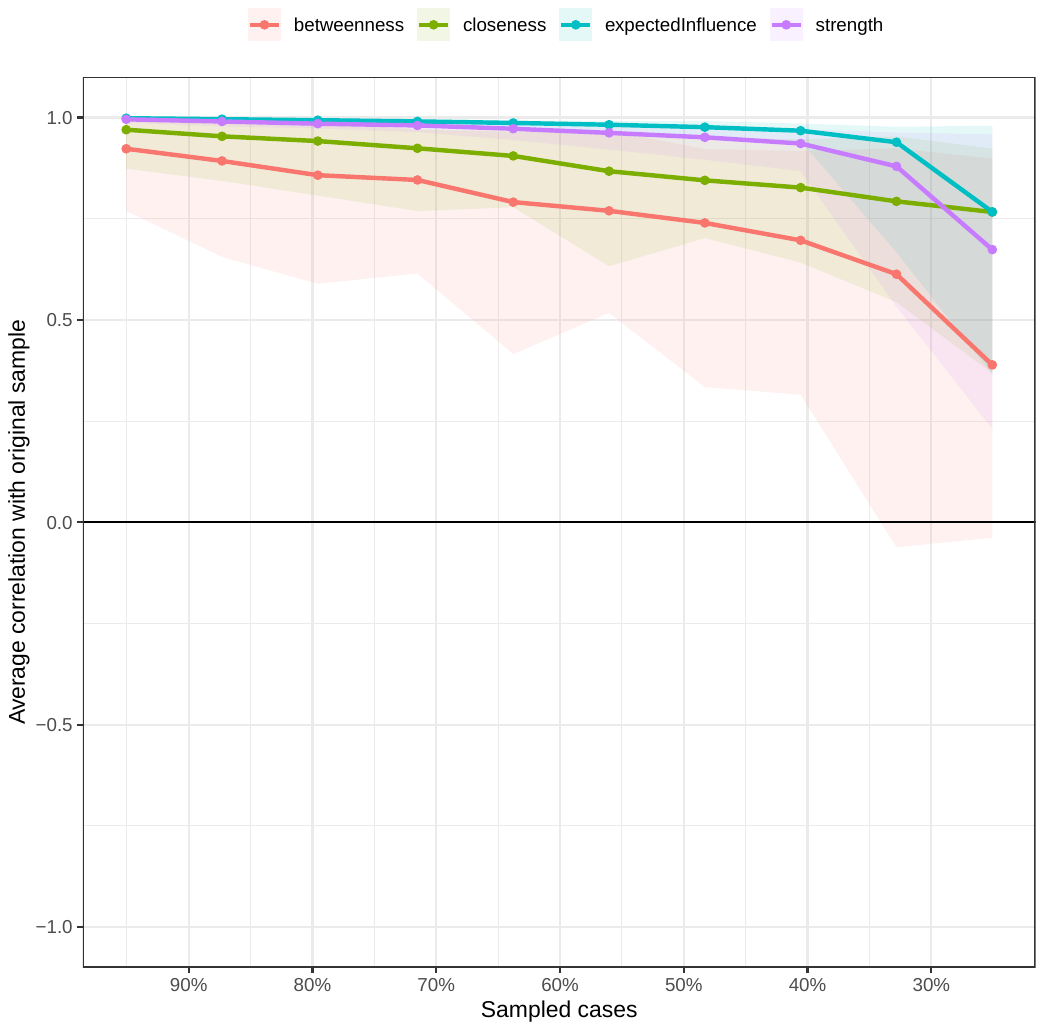 |

**Supplementary Fig. 5 Average correlations between centrality indices of the network estimated by the original sample and networks sampled with different propotions of cases.** Lines represent the means and areas represent the range from the 2.5th quantile to the 97.5th quantile. (A: Baseline with PHQ-9 sum scores; B: Month 12 with PHQ-9 sum scores; C: Baseline with PHQ-9 subscores; D: Month 12 with PHQ-9 subscores)

|  | Female | Male |
| --- | --- | --- |
| W0 | 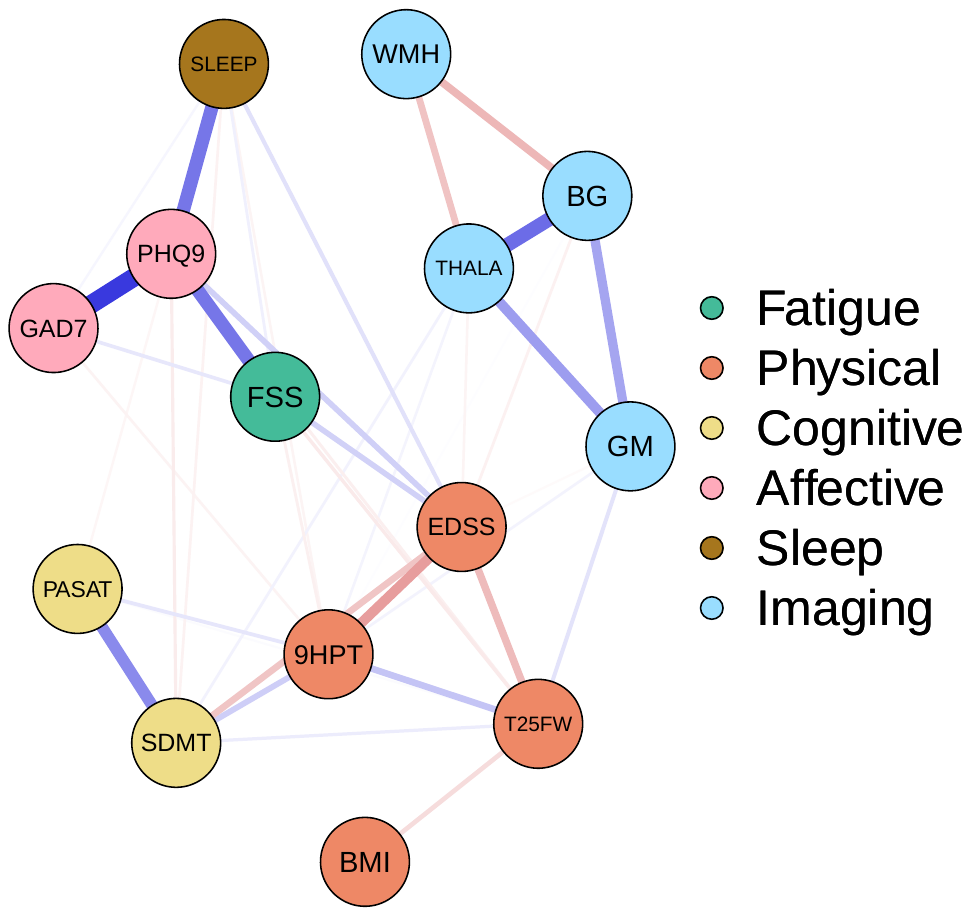  *n*=241 | 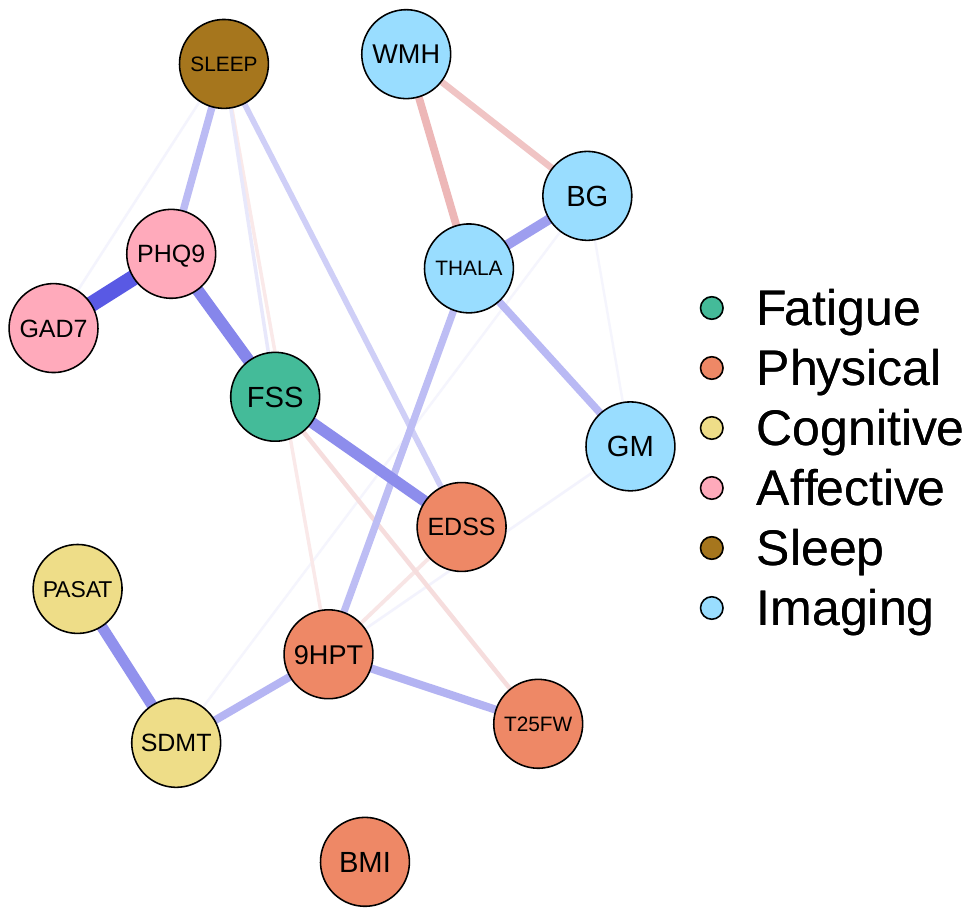  *n*=81 |
| W1 | 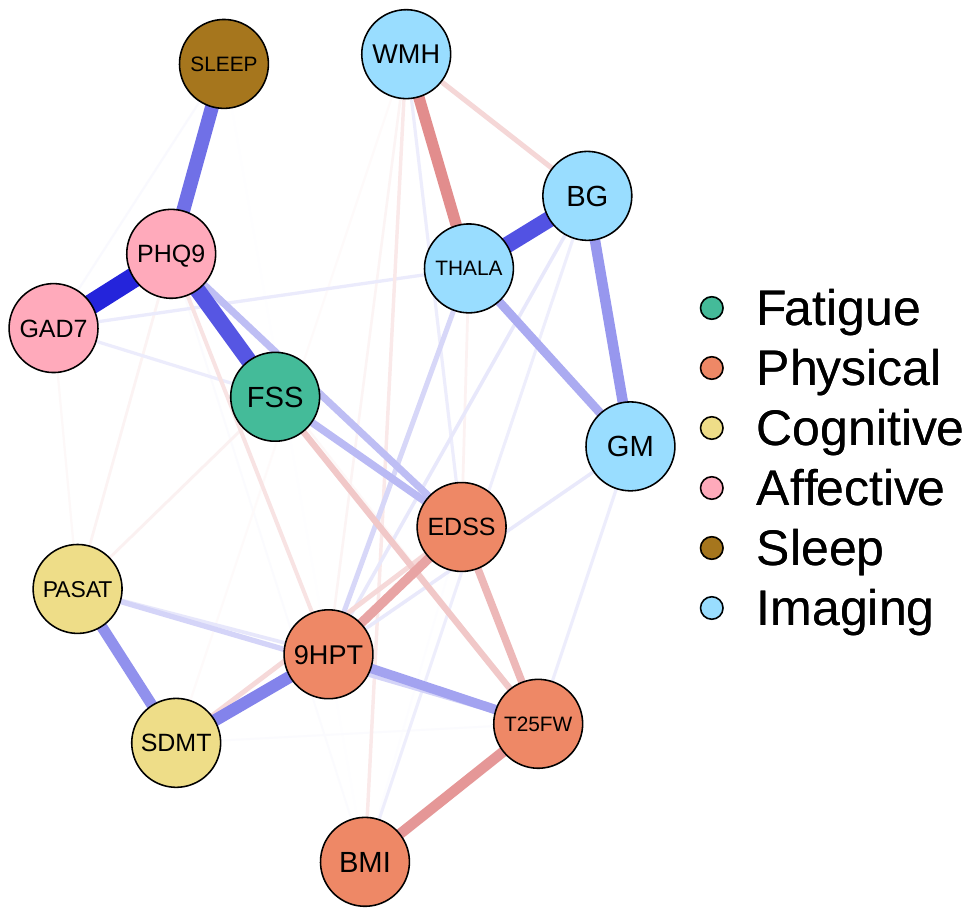  *n*=242 | 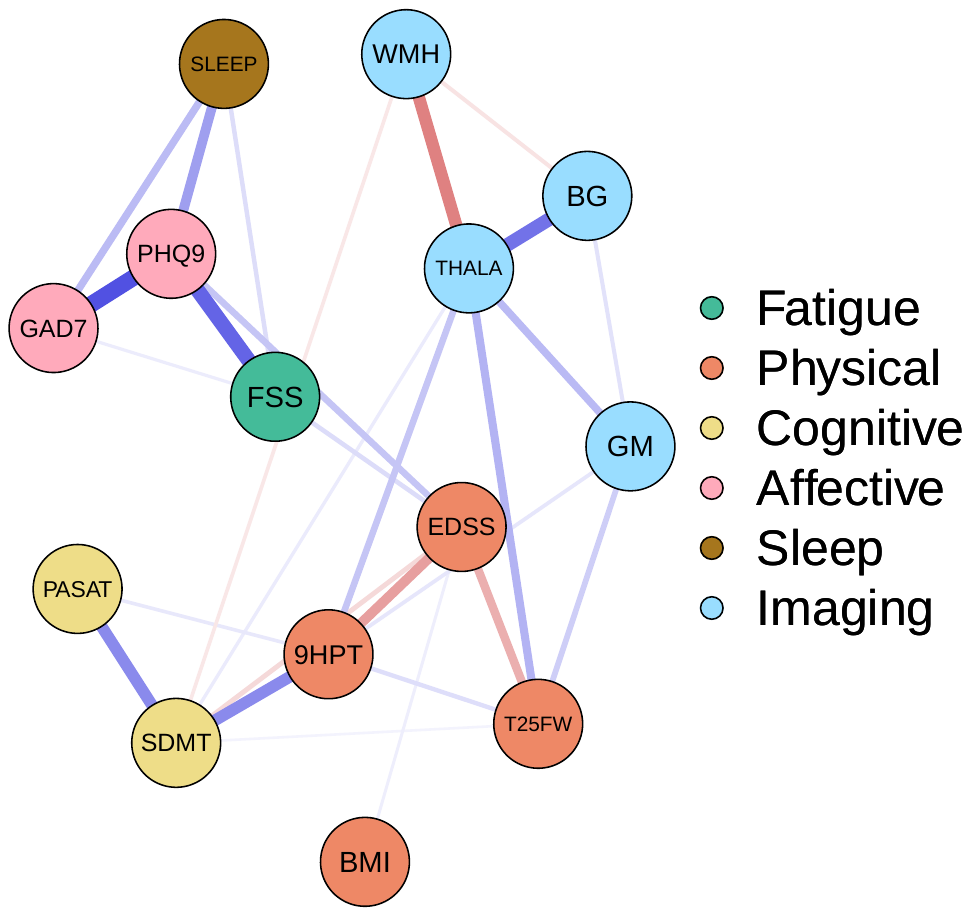  *n*=81 |

**Supplementary Fig. 6: Visualisation of the network estimation comparing different genders at baseline and month 12.** Blue edges indicate positive associations and red edges indicate negative associations. The width of the edges is proportional to the absolute value of the edge-weight. The colors of the nodes represent different domains.

Abbreviation: FSS=Fatigue Severity Scale; EDSS=Expanded Disability Status Scale; T25FW=Z score of Timed 25 Foot Walk test; BMI=Body Mass Index; 9HPT=Z score of Nine Hole Peg Test; PASAT=Z score of Paced Auditory Serial Addition Test; SDMT=Symbol Digit Modality Test; PHQ9=Patient Heath Questionnaire-9; GAD7=Generalized Anxiety Disorder-7 instrument; SLEEP = Multiple Sclerosis Impact Scale – Problems sleeping; WMH=Whole-brain white matter hyperintensity volume; cGM=cortical grey matter volume; BG=basal ganglia volume; THALA=thalamus volume

| **DMDs (Any type) *n*=235** | **DMDs (Never) *n*=88** |
| --- | --- |
| 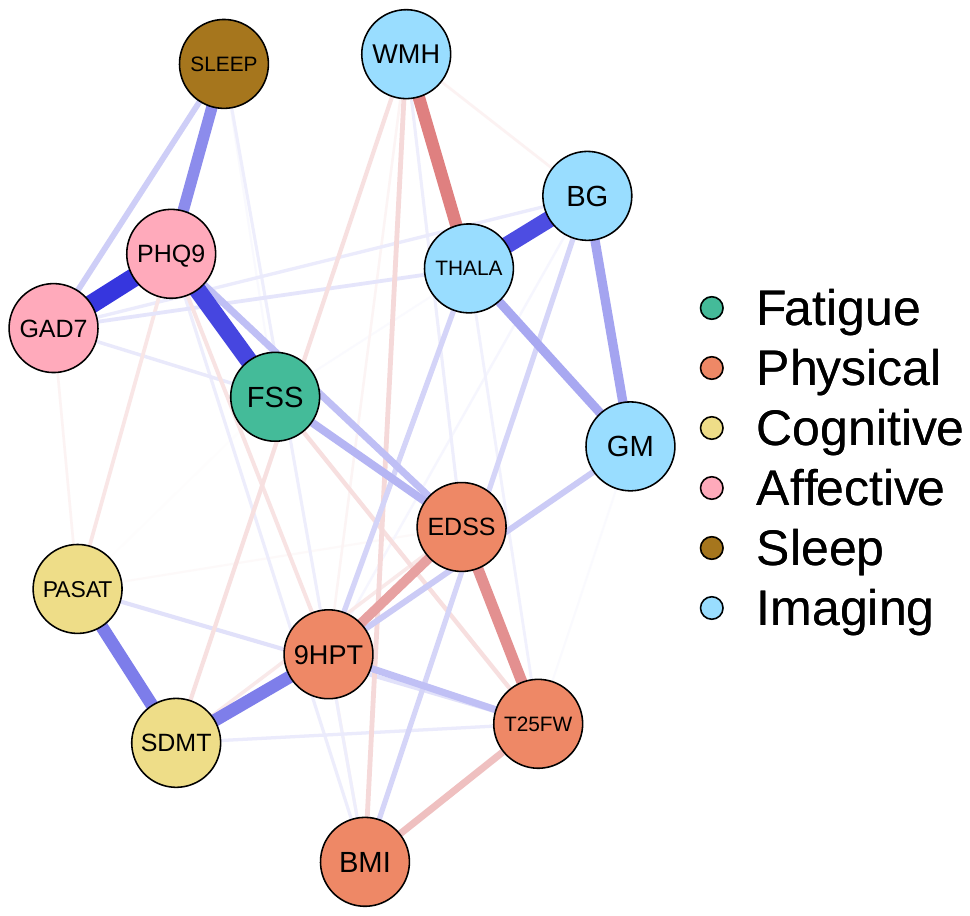 | 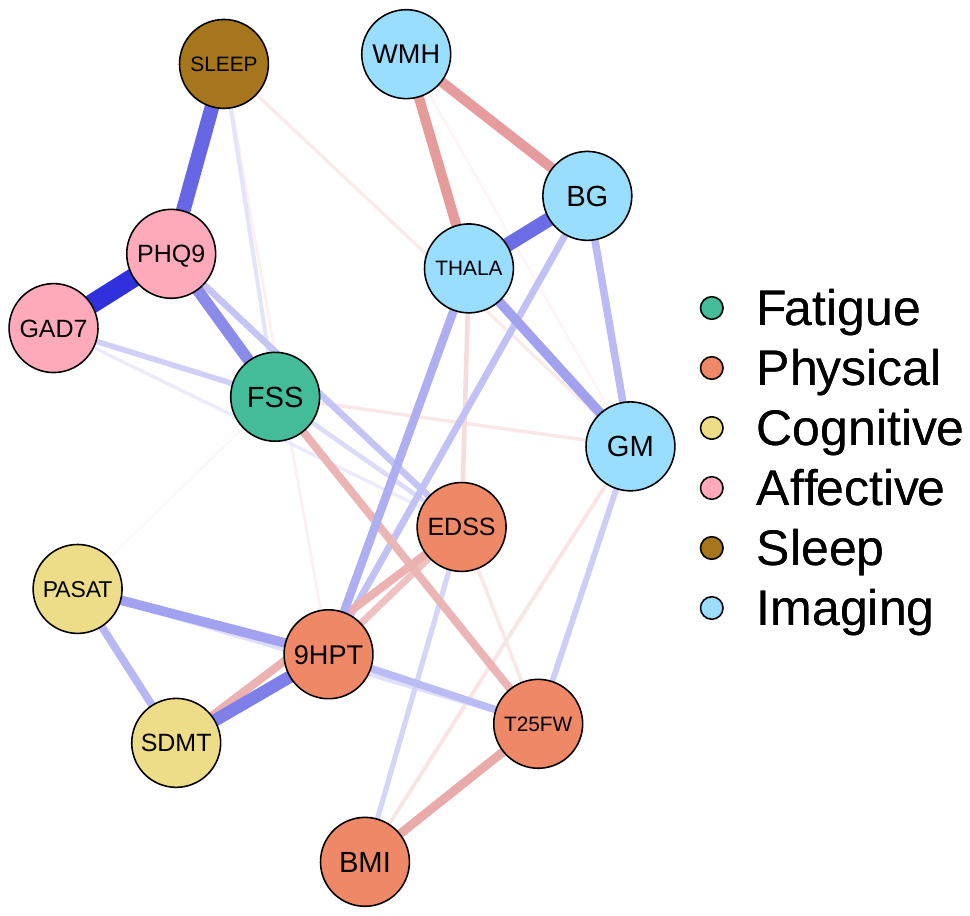 |

**Supplementary Fig. 7 Visualisation of the network estimations comparing those had used and had never used DMDs at month 12.** Blue edges indicate positive associations and red edges indicate negative associations. The width of the edges is proportional to the absolute value of the edge-weight. The colors of the nodes represent different domains.

Abbreviation: FSS=Fatigue Severity Scale; EDSS=Expanded Disability Status Scale; T25FW=Z score of Timed 25 Foot Walk test; BMI=Body Mass Index; 9HPT=Z score of Nine Hole Peg Test; PASAT=Z score of Paced Auditory Serial Addition Test; SDMT=Symbol Digit Modality Test; PHQ9=Patient Heath Questionnaire-9; GAD7=Generalized Anxiety Disorder-7 instrument; SLEEP = Multiple Sclerosis Impact Scale – Problems sleeping; WMH=Whole-brain white matter hyperintensity volume; cGM=cortical grey matter volume; BG=basal ganglia volume; THALA=thalamus volume

**Supplementary Table 1 Comparisons of clinical measures between subcohort eligible for network analysis and full cohort**

|  | Participants eligible for network analysis | | Full cohort | |
| --- | --- | --- | --- | --- |
|  | Baseline (*n* = 322) | Month 12 (*n* = 323) | Baseline (*n* = 440) | Month 12 (*n* = 392) |
| Female, No. (%) | 241 (74.8) | 242 (74.9) | 325 (73.9) | 292 (74.5) |
| Age at diagnosis, years, mean (SD) | 38.0 (10.2) | 37.8 (10.2) | 37.7 (10.2) | 37.9 (10.3) |
| BMI, mean (SD) | 27.6 (6.6) | 27.7 (6.8) | 27.8 (6.9) | 28.1 (6.9) |
| EDSS | 2 (1.5 – 3) | 2.5 (2 – 3) | 2 (1.5 – 3) | 2.5 (2 – 3) |
| zT25FW | 0.2 (-0.4 – 0.6) | 0.4 (-0.1 – 0.7) | 0.2 (-0.4 – 0.6) | 0.4 (-0.1 – 0.7) |
| z9-HPT | 0.2 (-0.6 – 0.7) | 0.4 (-0.4 – 1.00) | 0.1 (-0.6 – 0.7) | 0.4 (-0.4 – 1.00) |
| FSS | 34 (23 – 46) | 35 (19 – 50) | 35 (24 – 47) | 35 (20 – 50) |
| SDMT | 61 (54 – 68) | 62 (54 – 69) | 61 (54 – 68) | 62 (54 – 69) |
| PASAT | 47 (40 – 53) | 50 (42 – 55) | 46 (37 – 53) | 49 (40 – 55) |
| PHQ-9 | 6 (3 – 11) | 4 (1 – 8.5) | 7 (3 – 12) | 4 (2 – 9) |
| GAD-7 | 4 (2 – 7) | 3 (1 – 7) | 4 (2 – 8) | 4 (1 – 7) |
| Data above expressed as median (IQR) if not specified.  Abbreviation: BMI = Body Mass Index; EDSS = Expanded Disability Status Scale; zT25FW = Z score of Timed 25 Foot Walk test; z9-HPT = Z score of Nine Hole Peg Test; FSS = Fatigue Severity Scale; SDMT = Symbol Digit Modality Test; PASAT = Z score of Paced Auditory Serial Addition Test; PHQ9 = Patient Heath Questionnaire-9; GAD7 = Generalized Anxiety Disorder-7 | | | | |

**Supplementary Table 2A Correlation matrix at baseline with PHQ-9 sum scores**

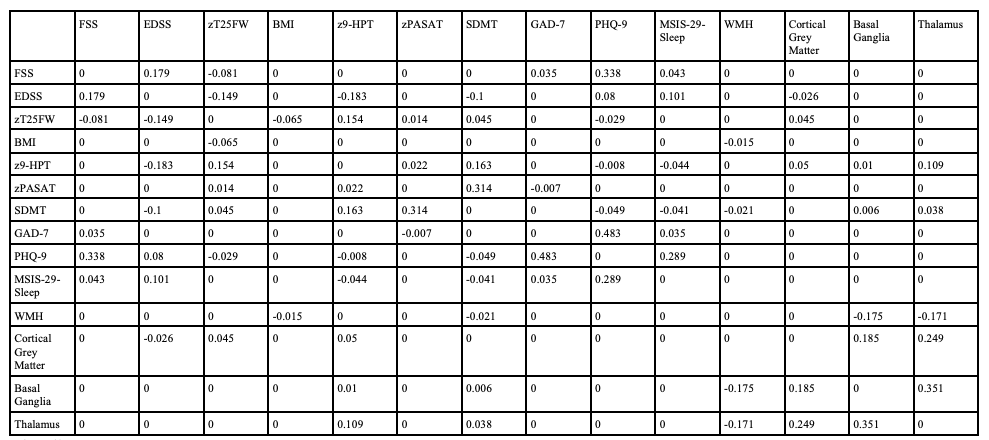

**Supplementary Table 2B Correlation matrix at month12 with PHQ-9 sum scores**

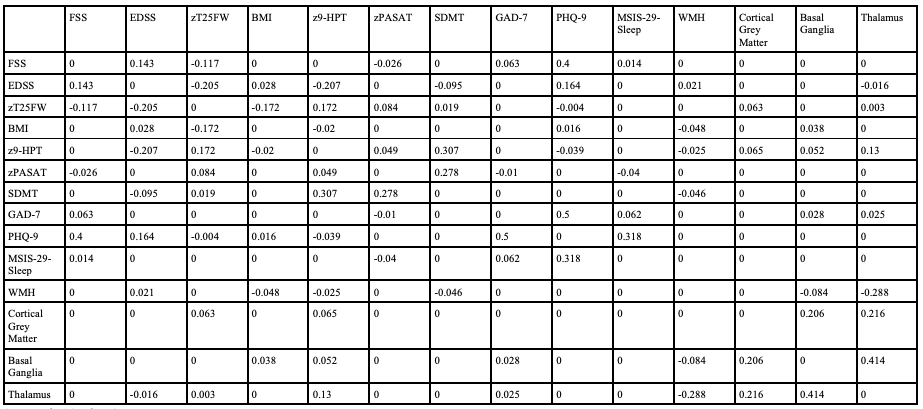

**Supplementary Table 2C Correlation matrix at baseline with PHQ-9 subscores**

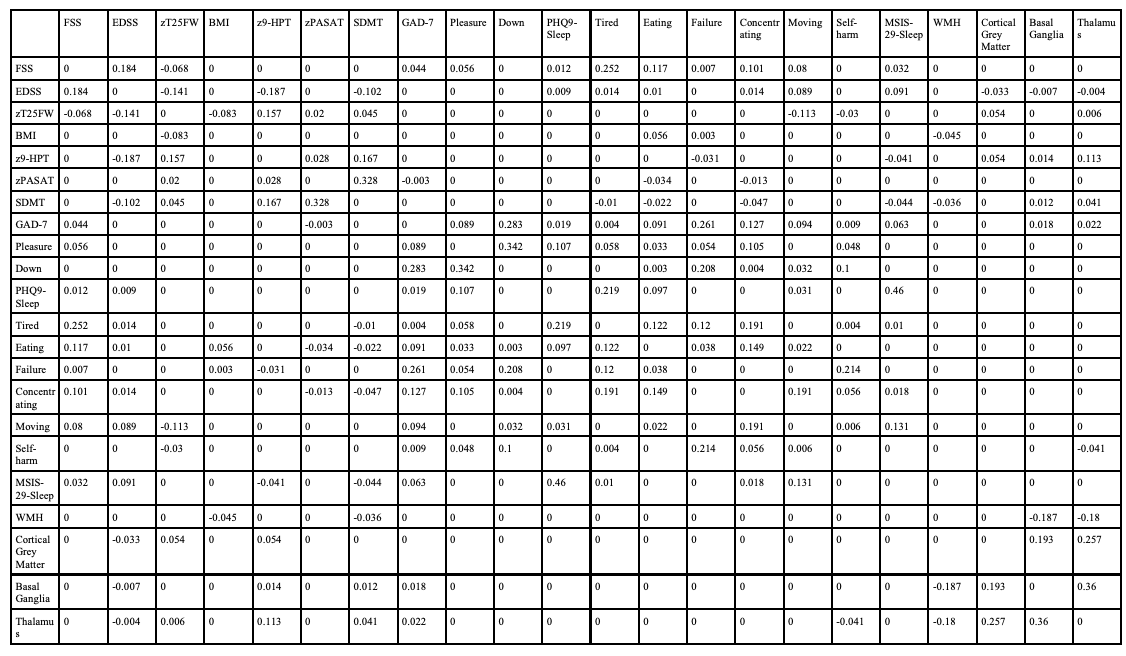

**Supplementary Table 2D Correlation matrix at month 12 with PHQ-9 subscores**

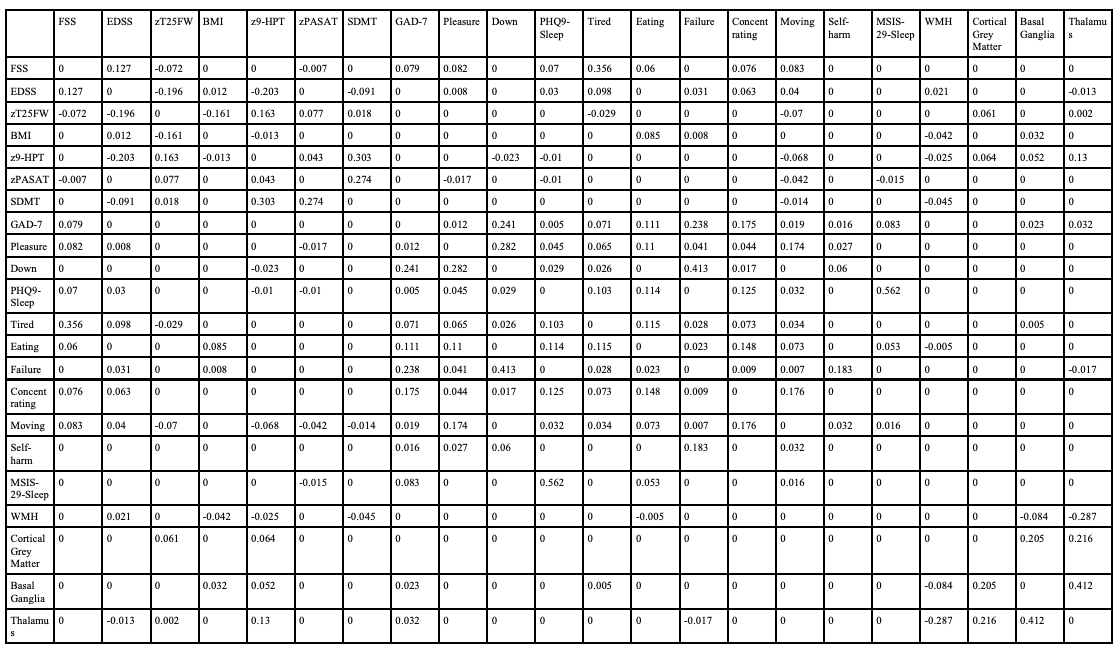

Supplementary Table 3: Correlation stability coefficients: Maximum proportions of dropped cases to retain correlation of 0.7 in at least 95% of the samples.

|  | PHQ-9 Sum Scores | | PHQ-9 Subscores | |
| --- | --- | --- | --- | --- |
|  | Baseline | Month 12 | Baseline | Month 12 |
| Strength | 0.593 | 0.672 | 0.516 | 0.672 |
| Closeness | 0.127 | 0.517 | 0.205 | 0.517 |
| Betweenness | 0.205 | 0.44 | 0.127 | 0.127 |
| Expected Influence | 0.671 | 0.749 | 0.593 | 0.594 |
